# Predicting multi-modal symptom trajectories across 7 years in veterans with chronic posttraumatic stress

**DOI:** 10.1101/2020.10.08.20185272

**Authors:** Benjamin Pierce, Thomas Kirsh, Erich Kummerfeld, Adam R. Ferguson, Thomas C. Neylan, Beth E. Cohen, Sisi Ma, Jessica L. Nielson

## Abstract

**Background:** Veterans are disproportionately affected by symptoms of post-traumatic stress (PTS) and associated poor health and psychosocial functioning. While most improve over time, others experience severe and persistent concerns. The ability to predict this latter group is critical for early intervention. Characterizing this subgroup has proven difficult, with most studies focusing on PTS and neglect a wider assessment of veterans’ wellbeing. Consequently, little is known about veterans who experience chronic symptoms and far-reaching impairment.

**Method:** The present study uses dimension reduction, growth mixture modeling, and clustering methods to identify veterans with the worst-faring trajectories of psychiatric symptoms, health, and psychosocial functioning, using data from the seven-year Mind Your Heart study (MYH) of people receiving Veterans Affairs services (n = 747). Random forest classification and feature selection were then used to examine predictors that distinguish the worst-fairing veteran group from others in the cohort.

**Results:** The combined analyses revealed a subgroup of veterans with severe and diverse symptoms across psychiatric domains, impairment in multiple facets of living, and poor health with deterioration over seven years. This subgroup was distinguished by transdiagnostic symptom severity and greater social isolation, avoidance, anhedonia, cynicism, anger/hostility, and immune response and inflammation.

**Conclusions:** Veterans whose distress spans multiple domains appear to be more broadly impaired, socially isolated, cynical or angry/hostile to others, and show elevated immunoreactivity and inflammation. Care for this population should be informed by a multidisciplinary approach that is conscious of veterans’ mental and physical health, and interpersonal needs.

## Introduction

Veterans are disproportionately affected by post-traumatic stress (PTS) and associated impacts on emotional, psychosocial, and physical health across the lifespan(1,2). Much research highlights the psychological and physical tolls of various military stressors and other traumatic events(3,4), with growing literature indicating this burden extends well beyond PTS into multiple facets of living(3,5). However, there are notable disparities in veteran outcomes across physical, emotional, and psychosocial domains that may be obscured by global statistics (4,6–13). While most veterans affected by PTS achieve symptom reduction over time and some improvement in functioning(8,10), a minority develop persistent symptoms with debilitating consequences(5,8).

Determining who fares worse among veterans presents challenges due to a breadth of factors potentially influencing a veterans’ trajectory over time. The high rates of co-morbidity(3), diverse psychosocial consequences(5,9,10), and varied health complications (5,9) among veterans suggest that a constellation of factors may differentiate veterans with poor outcomes from those who do better. Unfortunately, the majority of longitudinal research studies have emphasized PTS, while omitting other potentially salient outcome indicators. Consequently, while an array of polygenic factors and biomarkers (6), internal resilience and risk factors (7–9), peritrauma conditions (4,10,11) and experiences (14), and sociodemographic factors (12) may explain disparities in veteran outcomes, most of this evidence relies on characterizing veteran outcomes in a single domain related to symptoms of PTS.

Therefore, a multidimensional perspective is crucial to distinguish veterans with a chronic, pervasive profile of complaints from those with more limited or short-term symptoms. While previous research has developed predictive algorithms for relatively short-term PTS-related outcomes (14), there is very little information about what contributes to a chronic course of PTS with widespread mental and physical symptoms and psychosocial impacts. Given a wide assessment of outcome domains, current multivariate, data-driven approaches have the potential to derive combinations of variables that optimally distinguish veterans with the worst trajectories over time. Such approaches can provide important insights into the clinical features that most strongly distinguish veterans with the worst overall outcomes from those that are able to recover. Further, the distinctions made through these analyses can provide a clearly defined target for predictive analytics that may reveal novel predictors of prognostic course and salient therapeutic targets.

The present study aims to characterize veterans with the worst-faring features of psychiatric symptoms, health, and psychosocial trajectories through identifying multivariate dimensions and unobserved clusters that distinguish the varieties of prognosis in a 7-year longitudinal study of people receiving care through the VA system.

## Methods and Materials

### Participants

Participants included 747 patients recruited through the Mind Your Heart (MYH) Study (15) who were receiving care through the Department of Veterans Affairs medical centers in San Francisco and Palo Alto, California and reassessed each year. Participants were initially recruited between 2008 and 2010 using fliers and targeted mailings, and were excluded if their address was unstable or uncertain in the next 3 years. Detailed recruitment procedures have been previously published (15). The sample characteristics are displayed in **Table 1**. The sample was largely male and somewhat older, with about one-third meeting criteria for post-traumatic stress disorder at baseline.

**Table 1.** Sample demographics and diagnostic status at baseline.

| Variable | *N* (%) |
| --- | --- |
| <u>Demographics</u> |  |
| Age | 58.37 (11.30)<sup>a</sup> |
| Sex |  |
| Male | 703 (94.24%) |
| Female | 44 (5.90%) |
| Ethnicity<sup>b</sup> |  |
| Asian/Pac. Is. | 65 (8.71%) |
| Black/Af. Am. | 160 (21.45%) |
| Latinx/Latin Am. | 56 (7.51%) |
| White/Eur. Am. | 432 (57.91%) |
| Other than listed | 22 (2.95%) |
| Education |  |
| Less than H.S | 27 (3.62%) |
| H.S Graduate | 129 (17.29%) |
| Some College | 371 (49.73%) |
| College Deg. + | 218 (29.22%) |
| <u>12-Month DSM-IV Diagnosis<sup>c</sup></u> |  |
| Current PTSD | 258 (34.50%) |
| Major Depressive Disorder | 189 (25.30%) |
| Generalized Anxiety Disorder | 104 (13.92%) |
| Dysthymic Disorder | 46 (6.17%) |
| Bipolar-I | 41 (5.50%) |
| Bipolar-II | 26 (3.49%) |

<sup>a</sup> Age reported as *mean (standard deviation)*. <sup>b</sup> Listed categories for ethnicity included Asian/Pacific Islander; Black/African-American; Latinx, Hispanic, or Latin American; White/European American; or another ethnicity. <sup>c</sup> Diagnoses determined by structured clinical interview.

### Study Procedures

Participants completed extensive baseline assessments involving clinician-administered, self-report, and biometric measures (e.g., blood draw, electrophysiological, and treadmill stress-test measures). After baseline, participants were contacted yearly for seven years to complete brief assessments of mental and physical health outcomes. Full details on all measures in the study are provided on the MYH study website (15).

### Analytic Procedures

The present study aimed to characterize veterans’ wellbeing from a multidimensional lens and to identify predictors of disparities in veterans’ long-term trajectories across several psychiatric, functional, and psychosocial domains. Figure 1 presents the analytic workflow of the study. First, multivariate outcomes were derived using dimension reduction approaches to establish composite outcomes of distress and functioning in multiple domains. Next, multivariate and univariate approaches were used to stratify participants into better and worse-faring trajectories on each outcome domain individually, and on combinations of the domains together. Finally, Random forest classification and feature selection analyses were used to predict which participants had the worst-faring trajectories across the seven-year study period. Together, the dimension reduction, classification, and predictive analyses were able to highlight and describe participants in MYH showing greatest decline across outcome domains, as well as identify prognostic features that differentiated the worst-faring outcome trajectories from others. The reader is referred to the Supplementary Methods for a detailed description of all analytic procedures.

**Figure 1.**
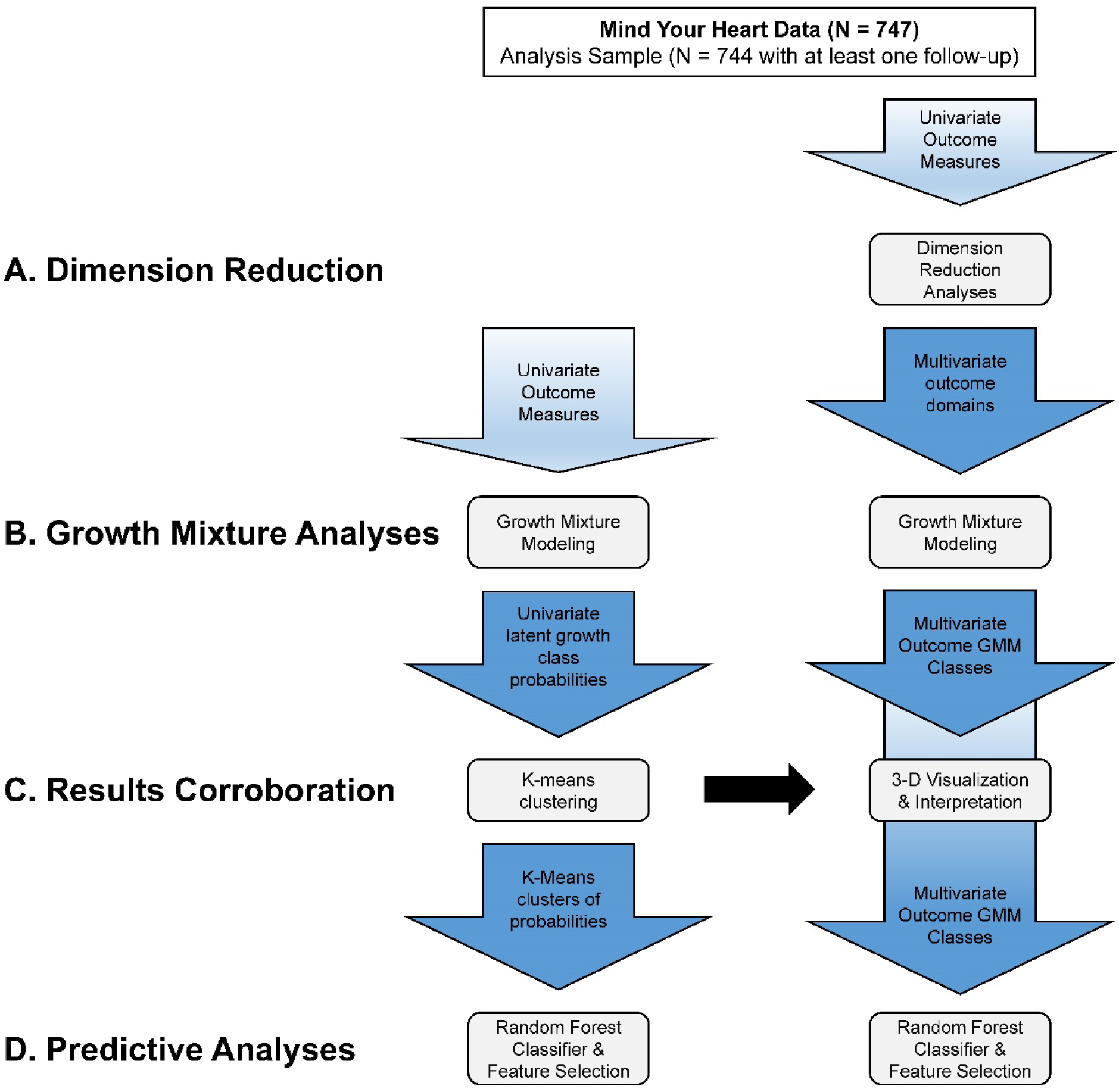
Study diagram showing analytic workflow and participant inclusion. GMM = Growth mixture modeling. Analyses performed on the univariate outcomes are represented by the vertical path on the left, and those performed on the multivariate outcomes are represented by the vertical path on the right. Vertical arrows reflect inputs to each stage of analysis. Horizontal arrows indicate integration across the results of the univariate and multivariate outcome analyses. The full dataset was reduced to N=744 to include only participants who had at least one follow-up visit. **A**. The first wave of analyses used principal components and confirmatory factor analyses to identify multivariate domains that represent interrelated group outcomes. **B**. Growth mixture analyses were performed on both the univariate and multivariate outcomes derived in the previous steps to differentiate participants with diverging trajectories on each univariate and multivariate outcome domain. **C**. K-means clustering was then used to reveal sub-groups of participants with high probabilities of showing different outcome trajectories on all univariate assessments. These participants were distinguished from others on the basis of their multivariate outcome scores at baseline. **D**. Random forest classification with feature selection was used to better understand categories of predictors for those in the worst-faring outcome trajectories.

### Univariate Measures

A range of univariate measures assessed yearly in MYH were incorporated in estimating the multivariate outcomes. These measures included self-report assessments of PTS (the PTSD Checklist-Military version(16)), depressive symptoms (the Patient Health Questionnaire(17)), physical functioning (the Short Form 36 – Physical scale(18–20)), absolute activity (past-week activity rating using a 5-point scale), comparative physical activity (past-week activity rating on a 5-point scale compared with one’s peers), health (the Short Form 36-Overall Health scale(21–23)), social functioning (the Short Form 36 – Social scale(21–23)), life quality (a 5-point life quality rating), and alcohol consumption (the Alcohol Use Disorders Identification Test – Consumption scale(24,25)) and were assessed annually after baseline assessment over a 7-year period.

### Multivariate Outcomes

Multidimensional outcome domains were identified using dimension reduction techniques that reduced the shared variance across univariate outcome measures to underlying dimensions or factors. These domains were established through applying nonlinear principal component analysis (NLPCA) (26) to the baseline univariate outcomes in randomized splits of the data to assess consistency in the underlying dimensions and loadings, and then re-running the NLPCA on the full dataset to obtain parameters for the full sample. Next, the stability of the variance dimensions across baseline and all seven yearly follow-ups was evaluated using confirmatory factor analysis (CFA) (27) to test equality constraints imposed on the dimensions and associated loadings across time. Once temporal stability of the dimensions and loadings was established, multivariate outcomes were computed as the linear combinations of univariate measures loading onto each dimension in the full-sample NLPCA. These analyses are described in-detail in the Dimension Reduction Analyses section of the Supplementary Methods. Code for these dimension reduction analyses can be found in the Supplemental Code for principal component analysis SPSS syntax (pages 1-7) and confirmatory factor analysis R syntax (pages 8-15).

### Latent Growth Mixture Models

Each multidimensional outcome was analyzed using growth mixture modeling to classify veterans into better or worse-faring trajectories. Specifically, latent growth mixture models (GMM) (28) were used to cluster participants based on their trajectories of change in the multivariate outcomes across the assessments. GMM uses maximum likelihood estimation procedures to identify classes of participants with statistically similar trajectories of change across-time. The trajectories associated with each latent growth class are assumed to characterize a subgroup of participants, with variations around each trajectory assumed to reflect ideographic deviations from a common pattern in that class. Each latent growth class trajectory is defined by average scores on the outcome (the intercept), the average rate of change in the outcome (the slope), and the shape of change across time (e.g., linear or polynomial). Participants were assigned to a latent growth class based on the correspondence between their individual trajectories and those identified through the GMM. The GMM analyses are described in-detail in the Growth Mixture Analyses section of the Supplementary Method. Code for GMM analyses can be found in the Supplemental Code for Python code for latent class growth analysis Mplus automation (pages 16-22).

### Cluster Analysis of Univariate Outcome Trajectories

The correspondence between the GMM classes on each multivariate outcome, and changes in the univariate outcomes over time, were used to corroborate which participants belonged to the worst-faring veteran group in order to compare these multivariate classifications with more traditional views of looking at univariate outcomes. GMMs were run on the univariate outcomes and the probabilities of participants being assigned to the poorest-faring classes on each univariate measure were obtained. Next, K-means clustering was used to group participants according to these probabilities, and their membership in these clusters was compared to their GMM class on each of the multivariate outcomes. In addition, the clusters were plotted along each of the multivariate outcome dimensions at baseline to assess correspondence between participants’ baseline multivariate scores and deterioration on groups of univariate outcomes over time. This provided a descriptive evaluation of whether faring poorly on the multivariate dimensions over time corresponded to deterioration across multiple univariate domains, and whether this deterioration was also associated with static multivariate outcome scores. The K-means clustering analyses are described in-detail in the K-Means Clustering section of the Supplementary Method. Code for performing K-means clustering can be found in the Supplemental Code (pages 23-26).

### Random Forest Classification Analysis

A random forest classifier with feature selection (RFC-FS) (29) was used to identify candidate predictors of membership in the worst-faring multivariate outcome class trajectory, as well as membership in the worst-faring k-means cluster based on the univariate outcome trajectories. Predictors were drawn sequentially across each measurement occasion to determine which measures showed sustained or emergent relations with the worst-faring trajectory over time. Feature selection was conducted by selecting the top features according to the random forest feature importance rankings and based on the area under the curve produced through including subsets of the top features. These predictors were then summarized based on their contents and relations to class membership to identify domains salient for veterans with the worst-faring multivariate trajectories. The RFC-FS analyses are described in the Random Forest Classification with Feature Selection Analysis section of the Supplementary Methods.

## Results

### Multivariate Outcomes

#### Multivariate outcome estimation

The NLPCA and CFA suggested three underlying dimensions that explained approximately 70.0% of the variance in the dataset, with each dimension explaining 44.4%, 14.1%, and 11.3% of the variance, respectively. The dimensions and associated loadings showed stability in the NLPCA across random splits of the data, as presented in Table 2. Further, the CFA factor loadings and residuals showed stability over time (Supplementary Table S1), with factor loadings consistent with the NLPCA as shown in Supplementary Figure S2. Therefore, the loadings from the full-sample NLPCA were used to compute weighted combinations of the univariate outcomes. The first linear combination reflected a composite measure of distress and functional impairment (Distress/Impairment outcome); the second was defined by higher activity levels concurrent with elevated emotional distress (Distress/Activity outcome); and the third primarily represented concerns captured by the alcohol consumption measure (Alcohol Use Concerns). Detailed results of these analyses are provided in the Principal Components Analysis and Confirmatory Factor Analysis sections of the Supplementary Results.

**Table 2.** Nonlinear PCA pattern matrices across splits of the sample and the full sample at baseline.

| Variable | First Split (n = 373) |  |  | Second Split (n = 374) |  |  | Full Sample (n = 747) |  |  |
| --- | --- | --- | --- | --- | --- | --- | --- | --- | --- |
|  | Component |  |  | Component |  |  | Component |  |  |
|  | 1 | 2 | 3 | 1 | 2 | 3 | 1 | 2 | 3 |
| AUD-C | -0.031 | -0.089 | 0.915 | 0.037 | 0.295 | 0.918 | -0.062 | -0.096 | 0.974 |
| GH | 0.759 | -0.265 | -0.227 | 0.755 | -0.045 | 0.143 | 0.755 | -0.170 | -0.049 |
| PCL | 0.752 | 0.507 | 0.084 | 0.731 | 0.432 | -0.193 | 0.743 | 0.484 | 0.041 |
| Absolute | -0.575 | 0.566 | -0.153 | -0.541 | 0.674 | -0.134 | -0.559 | 0.635 | -0.001 |
| Relative | -0.646 | 0.549 | -0.146 | -0.647 | 0.580 | -0.098 | -0.647 | 0.573 | -0.042 |
| PF | -0.661 | 0.114 | 0.314 | -0.574 | 0.226 | 0.055 | -0.624 | 0.147 | 0.206 |
| Quality | 0.758 | -0.070 | -0.067 | 0.691 | 0.241 | 0.176 | 0.727 | 0.060 | 0.043 |
| SR | -0.790 | -0.368 | -0.134 | -0.734 | -0.180 | 0.097 | -0.766 | -0.279 | -0.106 |
| PHQ | 0.814 | 0.412 | 0.133 | 0.779 | 0.357 | -0.199 | 0.795 | 0.405 | 0.070 |

Boxed loadings exceed .400. AUD-C = Audit – Consumption scale. GH = SF-36 General Health. PCL = PTSD Checklist. Absolute = Absolute activity. Relative = Relative activity. PF = Sf-36 Physical Functioning. Quality = Diminished life quality. SR = SF-36 Social Role. PHQ = Patient Health Questionnaire. Heat map represents PC loadings from -1 (blue) to 0 (white) to +1 (red).

#### Identifying multivariate outcome trajectories

Growth mixture models were estimated for each of the multivariate outcomes, and the resulting trajectories are presented in Figure 2 for the first two multivariate outcomes. For the Distress/Impairment outcome, four latent growth classes were identified, where the first class was differentiated by high and significantly increasing symptoms and impairment over time (Figure 2A), the second class by very low and stable symptoms and impairment (Figure 2B), the third class by moderate and significantly increasing distress and impairment (Figure 2C), and the fourth class by moderately-low and stable scores on this dimension (Figure 2D). For the Distress/Activity outcome, three latent growth classes were identified, none of which changed significantly over time, but were distinguished primarily by their different intercept values. The first class was characterized by low and stable scores (Figure 2E), the second by moderate and stable scores (Figure 2F), and the third by high and stable scores (Figure 2G). Finally, four latent growth classes were identified for Alcohol Use Concerns (Figure S3), with the first class defined by moderately-high and stable drinking (Figure S3A), the second and smallest class by especially high and stable alcohol use (Figure S3B), the third by low drinking (Figure S3C), and the fourth by modest use (Figure S3D). Across each of the outcomes, the largest classes tended to represent minimally symptomatic groups, whereas a small class represented participants showing deterioration or consistently elevated concerns. Detailed results of these analyses are described in the Growth Mixture Modeling section of the Supplementary Results and Table S2.

**Figure 2.**
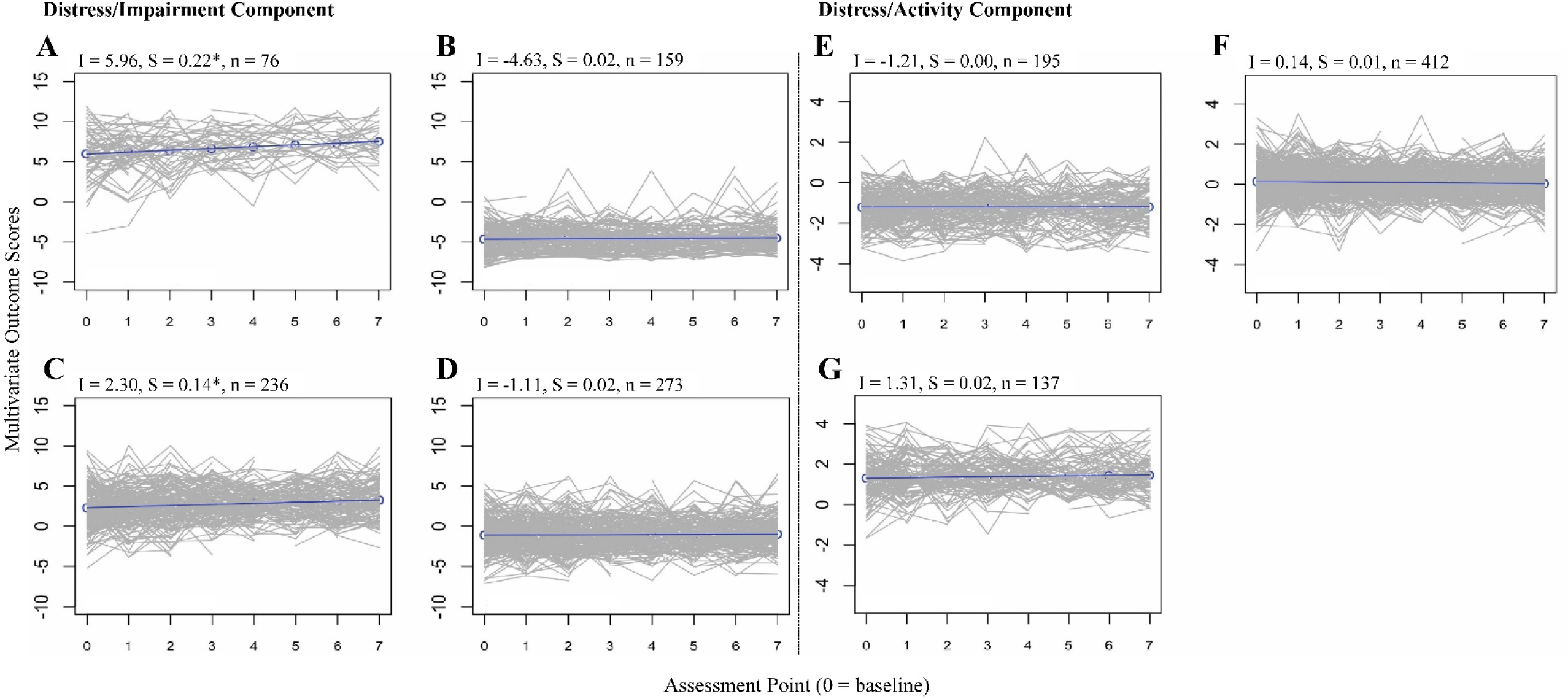
Latent growth class trajectories for the Distress/Impairment and Distress/Activity outcome dimensions. Three multivariate domains were identified based on 9 univariate outcome measures. The first dimension represents distress and impairment and was divided into four separate classes of patients, based on differing outcome growth trajectories within this dimension. These included **A**. high (scoring) and rising (increasing slope of blue population line), **B**. low (scoring) and stable (no change in slope of blue population line), **C**. moderate and rising, and **D**. slight and stable trajectories. The second dimension represents distress and activity and was divided into three separate clusters of patients based on outcome trajectories. These included E. low and stable, F. moderate and stable, and G. high and stable. The third dimension for alcohol use concerns are reported in Figure S3. Assessment points are measured in years. I = Intercept. S = Slope. n = Sample size for each class. *slope of the latent growth class is statistically significantly different from zero at *p* <.05. All class intercepts were statistically significantly different from zero.

### Univariate Outcomes

#### Identifying univariate outcome trajectories

GMMs were estimated for each of the univariate outcomes to classify their trajectories over time. Consistent with the multivariate GMMs, the classes estimated for each univariate outcome distinguished participants with a deteriorating or chronically-elevated trajectory. These are characterized fully in Table S2.

#### Clustering univariate outcome trajectories

Participants’ probabilities of being assigned to the worst-faring GMM class for each univariate outcome were clustered via k-means clustering. Six clusters provided the best solution across clustering indices (Figure S4), and one cluster reflected participants who fared poorly across all univariate outcomes, except for alcohol consumption. Table S3 provides the proportions of participants with each of the worst-faring univariate outcome trajectories by cluster. Details of these analyses are presented in the K-Means Clustering section and Figure S4 of the Supplementary Results.

#### Correspondence between Univariate and Multivariate Trajectories

All participants in the worst-faring cluster of univariate trajectories were in the worst-faring Distress/Impairment class, corroborating alignment between the multivariate and univariate analytic approaches. As displayed in Figure 3, scores on each of the multivariate outcome dimensions at baseline aligned with corresponding k-means clusters. The worst-faring cluster across univariate outcomes scored highly on the Distress/Impairment dimension (Figure 3A-3C); the cluster with elevated PTSD and depressive symptoms scored highly on the Distress/Activity dimension, while the cluster with elevated physical limitations had low scores on this dimension (Figure 3D-3F); and the cluster with elevated alcohol consumption scored highly on the Alcohol Use Concerns dimension (Figure 3G-3I).

**Figure 3.**
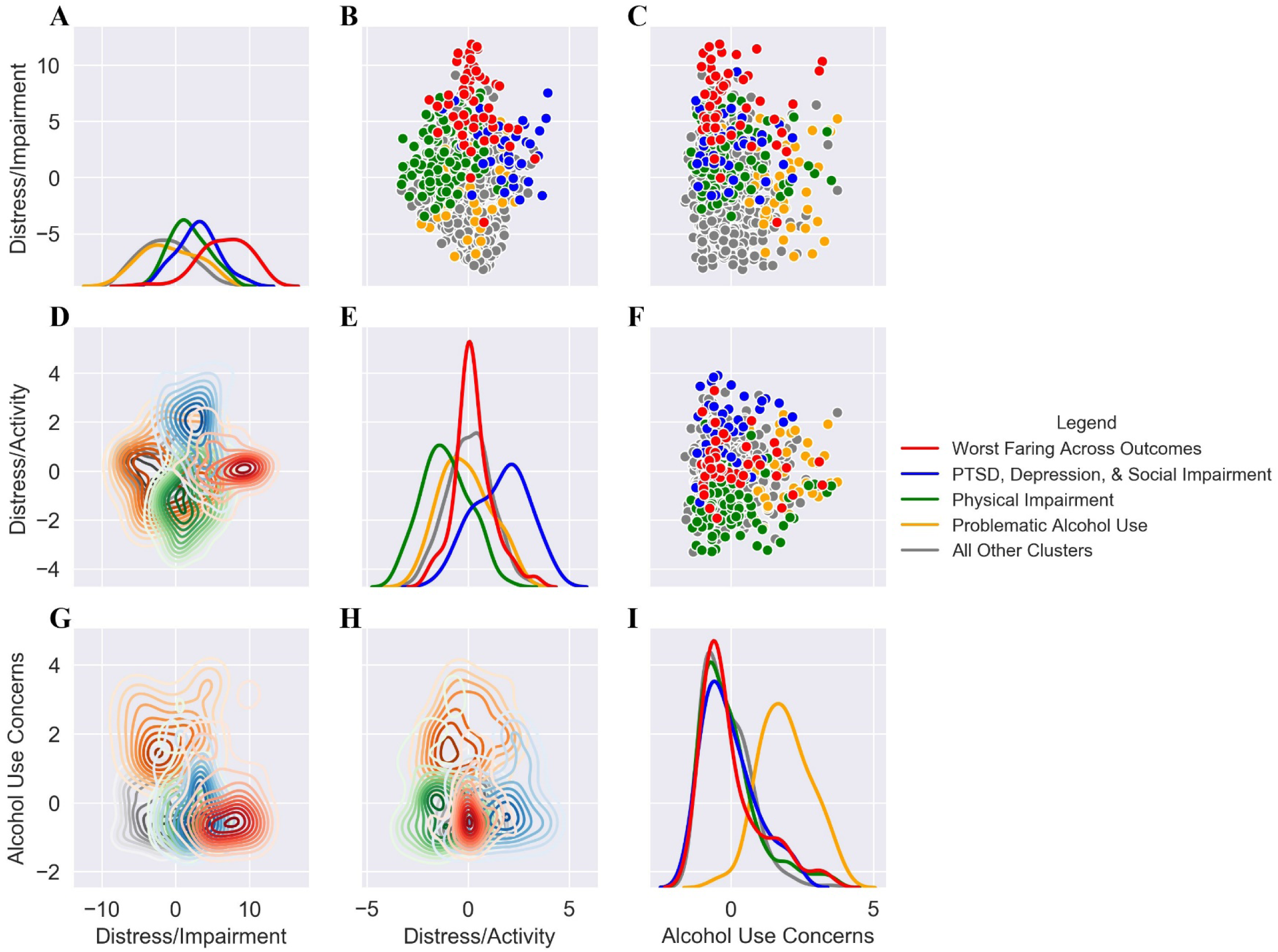
K-means cluster assignment, plotted by multivariate outcome dimension. Panels are arranged as a bivariate matrix of the three multivariate outcome domains identified by factor analysis and groups distinguished by color representing different k-means clusters derived from the univariate outcomes. Horizontal panels **A-C** represent the distress and impairment outcome, panels **D-F** represent the distress and activity outcome, and panels **G-I** represent the alcohol use concerns outcome. Panels **A, E**, and **I** show the univariate density plots for each multivariate outcome dimension, colored by k-means cluster. Panels **B, C**, and **F** show bivariate scatterplots among all pairs of multivariate outcomes colored by k-means cluster. Panels **D, G**, and **H** display these scatterplots plots as bivariate density diagrams, with each concentric line reflecting an increase in the density of the bivariate distribution within each cluster. Participants with a deteriorating course across univariate domains (red) are most strongly differentiated by baseline Distress/Impairment outcome scores. Participants faring worst in terms of the emotional and psychosocial concerns (blue) and participants faring worst in terms of physical complaints (green) are most strongly differentiated by scores on the Distress/Activity dimension at baseline. Finally, participants with the worst-faring alcohol consumption trajectory (yellow) are most strongly differentiated by baseline Alcohol Use Concerns. Altogether, baseline scores on the multivariate outcomes aligned strongly with participants’ k-means cluster assignment based on their univariate outcome trajectory probabilities.

#### Prediction of the Worst-Faring Outcome Groups

The RFC-FS analyses identified predictors with significant and stable associations with the worst-faring Distress/Impairment class and the k-means cluster reflecting participants with the poorest univariate outcome trajectories across domains. The top 20 features identified at each assessment point were selected for interpretation, as the areas under the curve when using the top 20 features at each assessment were comparable to those resulting from including more features in the analyses (Figure S5). Membership in both the worst-faring Distress/Impairment class and k-means cluster was commonly predicted by measures of social support, isolation, and impairment; overall depressive symptoms and specifically items assessing anhedonia (i.e., diminished interest or participation in activities); overall post-traumatic stress symptoms as well as specific avoidance and hyperarousal symptom clusters; hostility, cynical distrust, and anger/irritability measures; assessments of body weight and size; and indicators of immune activity and inflammatory response including Interleukins (IL)-10,IL-12, IL-6, and IL-1β as well as C-reactive protein. Measures in each of these domains emerged as salient predictors across at least half of the assessment occasions for both the worst-faring Distress/Impairment and k-means cluster groups. Table 3 presents a summary of the predictive domains represented by the top-20 predictive measures for the worst-faring distress/impairment class and worst-faring k-means cluster by assessment point.

**Table 3.** Salient predictive domains for the worst-faring distress/impairment class and k-means cluster. Marked cells represent domains identified in the top-20 predictive features of membership in the worst-faring group, as defined by either the multivariate distress/impairment class or the worst-faring k-means cluster of univariate trajectory probabilities. Each domain represents multiple variables and is described such that the domain is positively associated with the worst-faring group membership; for example, higher scores on variables in the social isolation/impairment domain are associated with a higher probability of belonging to the worst-faring class or cluster. Each assessment point includes all possible predictors from that assessment and all earlier assessments in the Mind Your Heart study in the random forest classifier. X’s represent salient predictive features for the worst-faring distress/impairment class, while #’s represent salient predictive features for the worst-faring k-means cluster of univariate trajectory probabilities. ^a^ Body size measures included BMI, waist-to-hip ratio, waist measurements, and weight in pounds or kilograms.

| Domain | Assessment Point |  |  |  |  |  |  |  |
| --- | --- | --- | --- | --- | --- | --- | --- | --- |
|  | B | 1 | 2 | 3 | 4 | 5 | 6 | 7 |
| Social Isolation/Impairment | X# | X# | X# | X# | X# | X# | X# | X# |
| Overall Depression | X# | X# | X# | # | X# | X# | X# | X# |
| Overall PTSD | X# | X# | X# | X# | X# | # | X# | X# |
| Avoidance | # | X# | # | # | X# | X# | X# | X# |
| Body size measures<sup>a</sup> | X# | X# | X# | # | X# | # | X# | # |
| Anhedonia | X# | X# |  | X# | # | X# | X# | # |
| Hyperarousal | X# | X | X# | # | # | # | # | X# |
| Cynicism/Hostility/Aggression |  | X# | # | X | X# | # | X# | # |
| Inflammatory / Immune Response | X | X |  | X# | X# | # | X# | # |

## Discussion

The present study used multiple, converging methods to characterize which veterans fare worst across psychiatric, health, and psychosocial functioning domains and to identify predictors of their prognosis. This data-driven investigation provides a multidimensional profile of who fares worse over time, and offers a snapshot of predictive features that may help distinguish veterans at risk for poor long-term outcomes. Analyses identified a group of veterans who showed a multidimensional profile of deterioration across PTS, depressive symptoms, social functioning, overall health, physical functioning, activity, and perceived life quality, and who were distinct from those showing a more limited profile of concerns or deterioration. The most broadly deteriorating group represented a minority of the total sample (10%) yet showed severe, stable symptoms and impairment in multiple domains, and was identified through the convergence of growth mixture analyses of multidimensional outcome trajectories and clustering on univariate outcome trajectories. This deteriorating group was distinguished from others by elevations across PTS and depressive symptoms; heightened avoidance, hyperarousal, and anhedonia; social isolation and low perceived social support; an interpersonal profile of heightened irritability, anger, and cynical mistrust of others; and elevated immune reactivity, inflammation, and body size. The present results echo prior research pointing to the involvement of a broad scope of emotional, psychosocial, and physiological systems in veterans’ wellbeing and suggest potential intervention targets within these systems that could alter the trajectories of the veterans who suffer most.

The characteristics of the worst-faring veteran group, along with factors predicting their 7-year trajectories, suggest this subset of veterans has widespread clinical needs, and may have limited success in treatments focused on just one or a few domains of concern. Rather, the most effective interventions for this population should be multifaceted and able to address the affective, interpersonal, and physical health-related dimensions of these veterans’ trajectories(30). Transdiagnostic features such as avoidance(31) and anhedonia(32) may be salient behavioral intervention targets given the co-occurrence of these symptoms identified in the worst-faring group. Conversely, tools from interpersonal psychotherapies may help this group of veteran’s process cynicism, mistrust, and feelings of hostility, and have previously shown promise in treating PTSD(33). Beyond structured therapy, strategies to enhance social support and involvement may provide a buffer against ongoing isolation, avoidance, and persistence of post-traumatic symptoms (5,34). Finally, the present findings suggest overactive inflammatory processes, physical impairment, and diminished overall health are important targets among veterans showing the greatest symptoms; inflammatory processes are closely related to post-traumatic responses(13), and behavioral health support along with rehabilitation may help this group achieve greater mobility and quality of life(35).

It is noteworthy that in addition to veterans with worst-faring trajectories, the present study identified additional groups of veterans with salient trajectories on more specific combinations of outcomes. The second multidimensional outcome revealed differences in veterans with primarily emotional health concerns versus those experiencing greater physical limitations, which were corroborated against trajectories on the univariate outcome domains. While prior research suggests a bi-directional relation between emotional distress and physical functioning among veterans(36), finding an orthogonal physical-emotional outcome dimension aligns with studies showing physical functioning is differentially related to clusters of PTSD symptoms(37) and that certain veterans remain physically active despite high levels of emotional and physical distress(38). As such, interventions addressing comorbidity among post-traumatic stress and depressive symptoms may be more salient for some veterans than behavioral health, or vice-versa, for veterans endorsing only emotional or physical complaints. Similarly, there was a distinction of alcohol use problems from other domains of concern that was also corroborated across univariate and multivariate outcome trajectories. Problematic alcohol use may follow a distinct trajectory from other outcomes and have unique prognostic predictors from those distinguishing the worst-faring overall trajectory. The independence of alcohol use from other outcomes may have emerged due to the older average age of the sample (39), or may reflect a more nuanced relation between alcohol use and other symptoms that could be moderated by problems with affect regulation (40) or contingent elevations in specific symptoms (e.g., hyperarousal, (41)). Further research is warranted to more explicitly test hypotheses comparing veterans with problematic drinking behavior with those showing other symptom profiles, identify mechanisms that heighten risk for problematic drinking in the presence of other symptoms, and clarify whether alcohol abuse and post-traumatic stress symptoms should be treated concurrently or sequentially. Together, these additional outcome dimensions highlight the need for effective triage to optimize intervention for veterans with more confined concerns, as well as the need to assess domain-specific risk factors, while also identifying those at risk for the most widespread distress and functional impairment.

The present data-driven, longitudinal investigation offers important insights into the question of “who fares worse?” in veteran populations through characterizing this population and candidate predictors of their trajectories over time. The study identified a veteran subgroup using the convergence of methods and measures, which may have otherwise been overlooked had each univariate measure been studied in isolation. The results highlight several important avenues for intervention and suggest the needs of veterans vary based on the severity of their distress and functional impairment. The findings point to the need for integrative perspectives on assessment and treatment that address symptom heterogeneity, transdiagnostic mechanisms of avoidance and anhedonia, the importance of social and behavioral health support, and relational patterns involving hostility and mistrust among the worst-faring veterans.

## Supporting information

Supplemental Methods, Results, and Code

## Data Availability

Data from the Mind Your Heart study is not publicly available, however details on the study design and data availability can be found at https://mindyourheartstudy.ucsf.edu/.

## Disclosures

### Data Access, Responsibility, and Analysis

BEC had full access to all the data in the study, and JLN had access to the de-identified dataset and takes responsibility for the integrity of the data and the accuracy of the data analysis.

