## Supplemental Methods, Results, and Code for "Predicting multi-modal symptom trajectories across 7 years in veterans with chronic posttraumatic stress"

**Table of Contents**

Supplementary Method……………………………………………………………….. 1

Supplementary Results……………………………………………………………...4

Supplementary References………………………………………………………… 13

Supplementary Code……………………………………………………………….. 14

Principal Components Analysis SPSS Syntax……………………………………..14

Confirmatory Factor Analysis R Syntax…………………………………………...18

Python Code for Latent Class Growth Analysis (LCGA) Mplus Automation…….24

R Code for K-Means Clustering of LCGA Class Probabilities……………………29

### Supplementary Methods

#### Dimension reduction analyses

The dimension reduction analyses were used to derive multivariate domains that accounted for covariance across the univariate outcome measures and were stable across random halves in the sample and across time. Nonlinear principal components analysis (NLPCA) [1] was used to identify the underlying dimensions associated with multiple outcome measures and corroborate these results across random halves of the sample. Confirmatory factor analysis (CFA) [2] was then used to assess the stability of these dimensions across all eight assessment points of the study.

##### Principal components analysis

The NLPCA was performed on two random splits of the data to assess for stability in the underlying component dimensions and loading pattern. NLPCA aims to maximize the relations between the univariate outcomes and the underlying principal components through permitting nonlinear associations between the outcomes and component dimensions. In NLPCA, the principal components are identified through eigenvalue decomposition after a discretization step in which variables are scaled for the analysis. Next, the principal components with eigenvalues greater than one were selected for interpretation. The proportions of variance accounted for in the outcomes by each principal component, and the correlations between each outcome and component (i.e., the loadings) are estimated. Each principal component is standardized to a variance of 1 and a mean of 0. Given stability in the dimensions and loadings across separate splits, the NLPCA was re-run using the full sample to obtain final parameter estimates based on the full data. Components were interpreted if their eigenvalues exceeded a 1.00, and loadings onto each component were interpreted if they exceeded a value of 0.400. The variables selected for interpretation on each component were then used to inform the substantive labeling of each component.

##### Confirmatory factor analysis

CFA was used to test the stability of the principal components over time. In CFA a hypothesized measurement model is specified through specifying latent factors onto which measures are expected to load (and to not load) as well as through the correlational structure among the measures and factors. This model is then tested against the observed covariance and mean structure of the data to assess the extent to which it fits or reproduces this structure. This is accomplished through iterative, maximum likelihood-based estimation that aims maximize the alignment between the model-estimated and the observed variances, covariances, and means among measures, given the constraints imposed by the latent measurement model. Based on the extent to which the measurement model fits the data, this model may be constrained or relaxed to improve the fit or test specific hypotheses about the configuration of the measures and latent factors. In the present analysis, we examined whether the loadings, intercepts, and residuals of the outcomes varied across time, based on a null hypothesis of measurement invariance across the eight measurement occasions. This analysis sought to replicate the findings of the NLPCA both in terms of the number of factors represented and in terms of the equality of model parameters over the eight assessment points.

The CFA's in the present analyses were parametrized as multi-group models and run at varying levels of model constraint. Each time-point was modeled as a distinct "group" of observations and constraints on the loadings, intercepts, and residuals associated with the measures were tested for equality across the time-point "groups." The likelihood of the observed data at each level of model constraint was compared with the likelihood of the data given the least constrained model to evaluate the extent to which constraints diminished the fit with the observed data. In addition, overall fit of the models at each level of constraint was assessed using the comparative fit index (CFI), root mean squared error of approximation (RMSEA), and squared root mean residual (SRMR) as relative fit indices[3]. In simulation studies, approximate criteria of a CFI of .95 or greater, an RMSEA of 0.08 or below, and an SRMR of below 0.08 have been suggested for adequate model fit. These fit indices were interpreted together given that the CFI and RMSEA are least affected by model estimation technique [4], while the SRMR provides an absolute assessment of fit based on discrepancies between the model-estimated and observed variance-covariance matrices [3].

Four levels of equality constraint were tested across the study assessment points. For the least-constrained model, only the configuration of measures relative to the factors was constrained across time-points, such that measures were set to load onto the factors if their NLPCA loadings exceeded a value of 0.4 for that factor. Next, equality constraints were imposed on the factor loadings to assess whether variation in the relations between the measures and factors across time exceeded a probability greater than chance. Third, equality constraints were imposed on both the loadings and residuals to assess of whether equal portions of variance in the outcome measures were expected to be accounted for by the factors and measure-specific residuals across time-points. Finally, if equality was retained across all other levels of constraint, equality was imposed on the loadings, intercepts, and residuals to test for strict invariance in the latent structure of these measures over time.

**Growth mixture analyses**

Growth mixture models (GMMs)[37] were used to identify latent classes of participants characterized by common intercept and slope values across assessment-points on each outcome. GMM's assume an invariant within-group slope and intercept parameter in each latent growth class, and classes are estimated by maximizing between-class variance in the intercept and slope parameters. The GMMs in the present study were run across ascending a-priori numbers of classes and used iterative, maximum likelihood estimation procedures to estimate the slope and intercept parameters associated with each class and determine the subjects' posterior probabilities of belonging in each class [37]. The models permitted the rate of change (the slope) and the intercepts of the growth trajectories to differ between classes for each outcome. The Bayesian information criterion (BIC) [38], Lo-Mendell-Rubin likelihood ratio Test (LMRT) [39], and bootstrap likelihood ratio test (BLRT) [38,40] were used to compare the fit of models with sequentially increasing numbers of classes, and the optimal number of classes was selected through a combination of minimizing the BIC, improvement in fit versus fewer classes as indicated by the BLRT and LMRT, and avoiding overfitting with exceedingly small class sizes.

The stability of the optimal class solutions identified for each outcome was assessed by running the same analysis across 100 random ninety-percent splits of the data. Stability was determined by the proportion of times the original class number was selected as optimal and based on likelihood ratio comparisons between the original model and each of the randomly selected splits. Given stability in the solution for the GMM, participants were assigned to a growth trajectory class based on their posterior probability of belonging in that class. Participants' class assignments on the multivariate outcomes were used as the outcome in the random forest classification predictive analyses, while participants' probabilities of being assigned to the worst-faring univariate outcome classes were clustered via k-means and the resulting clusters were entered into the random forest classification analyses.

**K-means clustering**

K-means clustering was applied to the results of the GMMs on the univariate outcomes to derive clusters for comparison against the multivariate growth classes. Specifically, k-means clustering was performed on participants' estimated probabilities of belonging to the worst-faring latent growth classes, which defined underlying clusters of participants whose probabilities shared proximity with *k* pre-defined centroids[7]. The k-means algorithm aims to find the cluster solution that minimizes within-cluster variability (i.e., based on the squared Euclidean distances of points in a cluster around that cluster's centroid) and maximizes between-cluster variability (i.e., based on the squared Euclidean distances between cluster centroids) such that participants are optimally distinguished between clusters and maximally similar within clusters. This algorithm uses an iterative procedure to identify the best solution for the locations of centroids for a given number of *k* clusters, wherein centroids are initially randomly placed and the Euclidean distances between points and the centroids are computed, then the location of the centroids is updated across iterations such that minimizes the within-cluster sum of squares[7]. In the present analysis, the optimal clustering solution for each value of *k* was determined through repeating this process 25 times with 100 iterations to minimize the within-cluster sum of squares for each repetition, and then selecting the optimal solution achieved across the 25 repetitions. The k-means clusters of univariate trajectory probabilities were then compared to the multivariate outcome scores and growth trajectory classes to corroborate which participants fell into the worst-faring group across outcomes.

**Random forest classification and feature selection analysis**

Random forest classification analyses with feature selection (RFC-FS) [6] was used to identify candidate predictors of which participants belonged to the worst-faring multivariate GMM trajectories as well as which participants belonged to the worst-faring k-means cluster. Each RFC-FS was run with a binary dependent variable indicating whether the participant was assigned to the worst-faring growth class or cluster compared with all other classes or clusters. RFC-FS involves growing multiple decision trees based on prediction of class membership through recursive partitioning of the sample, cross-validating that decision tree across subsets of the data, and selecting the variables that most strongly distinguish among participants assigned and not assigned to the class. The recursive partitioning algorithm entails selecting the variable at each node (i.e., branching point) of the decision tree that maximally distinguishes participants assigned to the class from those not assigned, and then repeating this process until no further partitions can be performed. These decision trees was cross validated through repeating 5-fold cross-validation across 10 iterations of folding the data (i.e., 10 x 5 fold cross-validation) [42]. Next, different subsets of features ranging from the top one to top 50 features were examined to identify the optimal number of predictors for distinguishing the worst-faring class or cluster of participants based on their sensitivity and specificity (i.e., using area under the curve)[43]. The best features were assessed sequentially at each measurement occasion by first including only measures assessed at baseline and then adding measures from each year of the study up to year 7. The top 20 features showing stability across time, or which emerged and remained salient following a given measurement occasion, were then interpreted as candidate prognostic predictors for the worst-faring group of veterans.

#### Supplementary Results

##### Principal components analysis results

Three primary components were identified in each split of the data as well as when the analysis was applied to the full dataset. In the first split, the first, second, and third principal components explained 46.54%, 14.23%, and 12.00% of the variance across outcomes, respectively. In the second split, the first, second, and third principal components explained 41.90%, 14.84%, and 11.24% of the variance across outcomes. For the full dataset, the first, second, and third principal components explained 44.38%, 14.12%, and 11.28% of the variance across outcomes. Loadings estimated from the NLPCA are presented in the main text in Table 2.

##### Confirmatory factor analysis results

Based on the results of the NLPCA, an initial CFA model was specified with two orthogonal factors and a unique dimension associated with the AUD-C. Figure S2 displays the hypothesized loading path diagram and residual structure across Distress/Impairment, Distress/Activity, and Alcohol Use Concerns dimensions for the CFA. In addition to what is displayed, the mean structure of the items was estimated freely, contributing an additional 25 degrees of freedom at each time-point across intercepts (for the continuously-scaled measures) and thresholds (for ordinally-scaled measures). The fit of this least-constrained CFA model was adequate according to global fit indices (CFI = .985, RMSEA = .074, and SRMR = .068). Given this, equality constraints were subsequently evaluated based on the chi-square likelihood ratio test statistic by comparing the more-constrained models to this least-constrained parametrization.

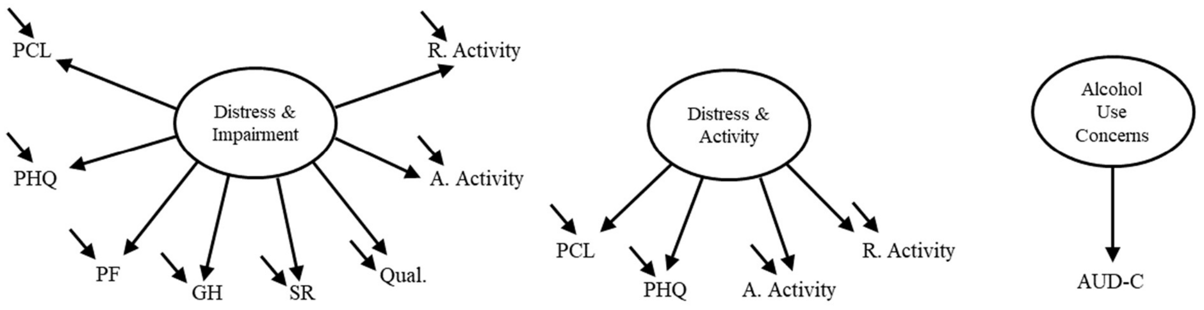

**Figure S1. Diagram of hypothesized measurement model.** AUD-C = Audit – Consumption scale. GH = SF-36 General Health. PCL = PTSD Checklist. Absolute = Absolute activity. Relative = Relative activity. PF = Sf-36 Physical Functioning. Quality = Diminished life quality. SR = SF-36 Social Role. PHQ = Patient Health Questionnaire. Arrows running from the factors to the measures represent hypothesized loadings. Small arrows represent measure-specific residuals.

The results of the chi-square likelihood ratio tests comparing model constraints against the least constrained model are presented in Table S1. Significant chi-square test statistics indicate the null hypothesis of equality across time-points was rejected, such that there was a difference in fit between the unconstrained model and the model given a set of equality constraints. As this table shows, equality of loadings and equality of both loading and residuals was retained across time-points. However, equality of the intercepts across measurement occasions was rejected based on the chi-square likelihood ratio test, suggesting participants' average values on the individual measures likely differed.

**Table S1. Chi-square likelihood ratio tests comparing each level of equality constraint across time-points with the least-constrained model**

| Type of constraint | $\chi^2$ Likelihood Ratio Test |
|---|---|
| Equality of loadings | $\chi^2(84) = 98.74$<br>$p = .130$ |
| Equality of loadings & intercepts | $\chi^2(210) = 293.06$<br>$p < .001$ |
| Equality of loadings & residuals | $\chi^2(112) = 130.57$<br>$p = .111$ |
| Equality of loadings, intercepts, & residuals. | Not tested because equality of the intercepts was rejected. |

Based on the results of the chi-square likelihood ratio tests, the global fit indices and parameters estimated from the model including equality of loadings and residuals across time-points were interpreted. This model implied equal variance-covariance matrices among the observed measures over time and equal variances of the latent variables across time-points, while allowing the mean structure of the measures to vary. Based on the global fit indices, this constrained model fit the data well (CFI = .979, RMSEA = 0.071, SRMR = .083). The results of the CFA's thus lend support for the stability of the factors across time-points. The present findings suggest the variance-covariance matrix among the measures can be adequately reproduced based on a measurement model including one Distress/Impairment factor with loadings from all outcome measures except the AUD-C; a Distress/Activity factor with loadings onto the PHQ, PCL, absolute activity, and relative activity variables; and a third orthogonal Alcohol Use dimension represented by the AUD-C. These results also suggest that the mean structure among the items varied over time, indicating that while the patterns of variance-covariance observed at each time-point may have been similar, there were differences in participants' relative levels of reporting on each of the outcomes over time. The standardized loadings from the final CFA model are presented in comparison to the NLPCA loadings based on full sample in Figure S2. As this figure illustrates, the loadings were largely similar across modeling approaches; slight variations in loadings may reflect differences in their meaning across models, where NLPCA loadings reflect variance explained in the component by the items and CFA loadings reflect variance explained in the items by the factors.

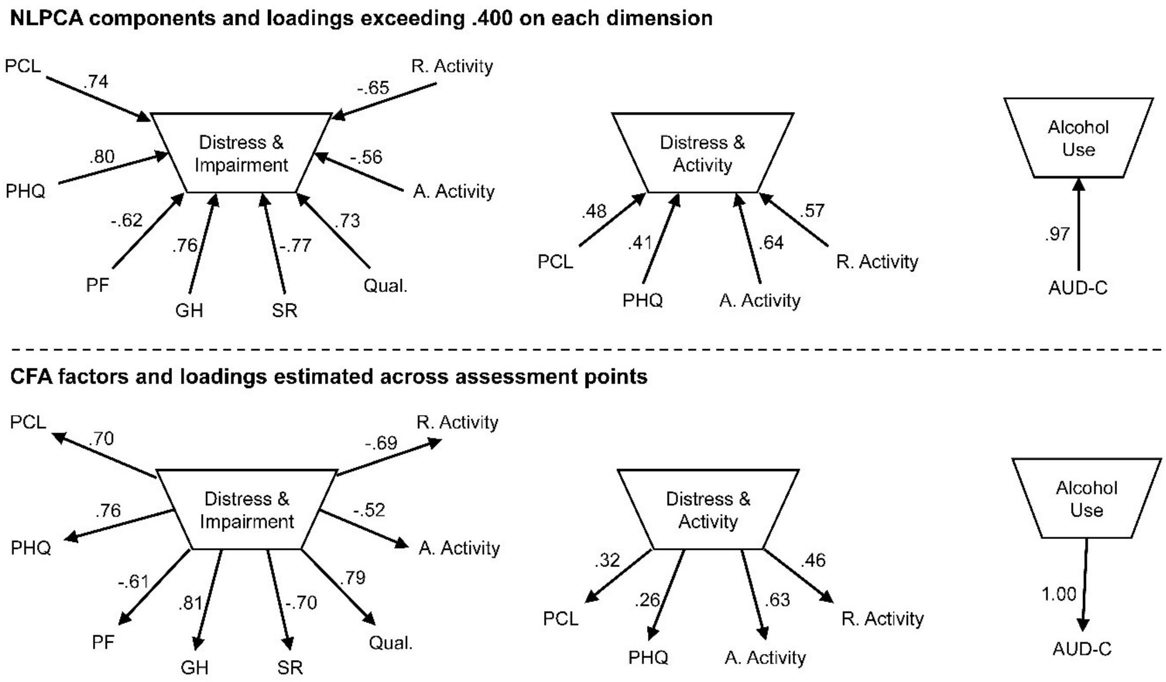

**Figure S2. Dimensions and associated loadings from the NLPCA and CFA analyses.** NLPCA = Nonlinear principal components analysis. CFA = Confirmatory factor analysis. AUD-C = Audit – Consumption scale. GH = SF-36 General Health. PCL = PTSD Checklist. Absolute = Absolute activity. Relative = Relative activity. PF = Sf-36 Physical Functioning. Quality = Diminished life quality. SR = SF-36 Social Role. PHQ = Patient Health Questionnaire.

**Growth mixture modeling (GMM) results**

The slope and intercept values associated with each of the GMMs are presented in Table S2. Figure 2 of the main manuscript supplement shows the latent growth trajectories associated with each class estimated for the Distress/Impairment and Distress/Activity multivariate outcomes. The latent growth trajectory associated with the Alcohol Use dimension is presented in Figure S3 of this supplement. The worst-faring Distress/Impairment class was distinguished by a profile of rising severity, whereas the worst-faring Distress/Activity and Alcohol Use Concerns classes were distinguished by sustained elevations. The GMMs on each univariate outcome paralleled the pattern of results found across the multivariate outcome domains; there tended to be larger classes of participants with fewer symptom complaints or functional concerns and a smaller class representing a subgroup of veterans with a more persistent or deteriorating course on a given outcome measure.

**Table S2. Latent growth class characteristics**

| **Measure**<br>**Class** | **Class Size** | **Slope** | **Intercept** |
|---|---|---|---|
| Multivariate Outcome Measures | | | |
| Distress/Impairment | | | |
| Class 1 – high & rising distress/impairment | 76 | 0.22* | 5.96 |
| Class 2 – low & stable distress/impairment | 159 | 0.02 | -4.63 |
| Class 3 – moderate & rising distress/impairment | 236 | 0.14** | 2.30 |
| Class 4 – slight & stable distress/impairment | 273 | 0.02 | -1.11 |

| | | | |
|---|---|---|---|
| Distress/Activity | | | |
| Class 1 – low & stable symptoms/activity | 195 | 0.00 | -1.21 |
| Class 2 – moderate & stable symptoms/activity | 412 | -0.01 | 0.14 |
| Class 3 – high & stable symptoms/activity | 137 | 0.02 | 1.31 |
| Alcohol Use Concerns | | | |
| Class 1 - moderate & stable concerns | 98 | 0.03 | 1.08 |
| Class 2 – high & stable concerns | 29 | 0.03 | 2.59 |
| Class 3 – very low & stable concerns | 373 | -0.01 | -0.66 |
| Class 4 – low & stable concerns | 244 | -0.01 | 0.29 |
| <u>Univariate Outcome Measures</u> | | | |
| PTSD Checklist | | | |
| Class 1 - moderate & stable symptoms | 138 | -0.30 | 44.51 |
| Class 2 - high & declining symptoms | 143 | -0.97** | 58.59 |
| Class 3 - mild & declining symptoms | 143 | -0.47* | 33.61 |
| Class 4 - high & stable symptoms | 93 | -0.08 | 69.05 |
| Class 5 - very low & declining symptoms | 229 | -0.33* | 22.49 |
| Patient Health Questionnaire | | | |
| Class 1 - mild & stable symptoms | 203 | -0.15 | 8.69 |
| Class 2 - moderate & stable symptoms | 130 | -0.07 | 13.64 |
| Class 3 – high & increasing symptoms | 68 | 0.37* | 17.94 |
| Class 4 - low & declining symptoms | 344 | -0.15* | 2.93 |
| Social Functioning | | | |
| Class 1 - low & stable functioning | 178 | -0.03 | 3.03 |
| Class 2 - very low & stable functioning | 94 | -0.04 | 2.02 |
| Class 3 - high & increasing functioning | 222 | 0.08** | 3.51 |
| Class 4 - very high & rising functioning | 252 | 0.03* | 4.53 |
| Overall Health | | | |
| Class 1 - very poor & declining health | 62 | 0.10** | 4.10 |
| Class 2 - poor & declining health | 218 | 0.07** | 3.42 |
| Class 3 - fair & slightly declining | 243 | 0.03* | 2.78 |
| Class 4 - positive & improving health | 79 | -0.03* | 1.42 |
| Class 5 - positive & stable health | 145 | -0.02 | 2.11 |
| Quality of Life | | | |
| Class 1 - positive & stable life quality | 168 | -0.05 | 2.36 |
| Class 2 - positive & rising life quality | 107 | -0.11** | 1.63 |
| Class 3 - fair & stable life quality | 280 | 0.04 | 2.94 |
| Class 4 - poor & declining life quality | 192 | 0.19** | 3.64 |
| Physical Functioning | | | |
| Class 1 - fair & stable functioning | 166 | -0.01 | 24.61 |

| | | | |
|---|---|---|---|
| Class 2 - high & declining functioning | 72 | -1.17** | 25.39 |
| Class 3 - low & declining functioning | 47 | -0.76** | 17.53 |
| Class 4 - low & stable functioning | 112 | 0.09 | 18.66 |
| Class 5 - high & stable functioning | 349 | 0.04 | 28.22 |
| Absolute Activity | | | |
| Class 1 - moderate & increasing activity | 253 | 0.07* | 2.42 |
| Class 2 - low & stable activity | 146 | -0.02 | 1.18 |
| Class 3 - high & declining activity | 32 | -0.44** | 3.94 |
| Class 4 - high & slightly increasing activity | 315 | 0.03* | 3.93 |
| Relative Activity | | | |
| Class 1 - moderate & stable activity | 251 | -0.01 | 2.75 |
| Class 2 - low & stable activity | 119 | -0.02 | 1.82 |
| Class 3 - moderately high & stable | 232 | 0.01 | 3.63 |
| Class 4 high & slightly increasing | 144 | 0.03* | 4.46 |
| AUD-Consumption | | | |
| Class 1 - low & declining alcohol use | 432 | -0.03* | 0.85 |
| Class 2 - high & stable alcohol use | 58 | 0.02 | 7.87 |
| Class 3 - moderate & stable alcohol use | 256 | -0.02 | 3.87 |

*Note.* *$p < .05$. **$p < .01$. *All intercepts were statistically significantly different from 0. AUD = Alcohol Use Disorders Identification Test.*

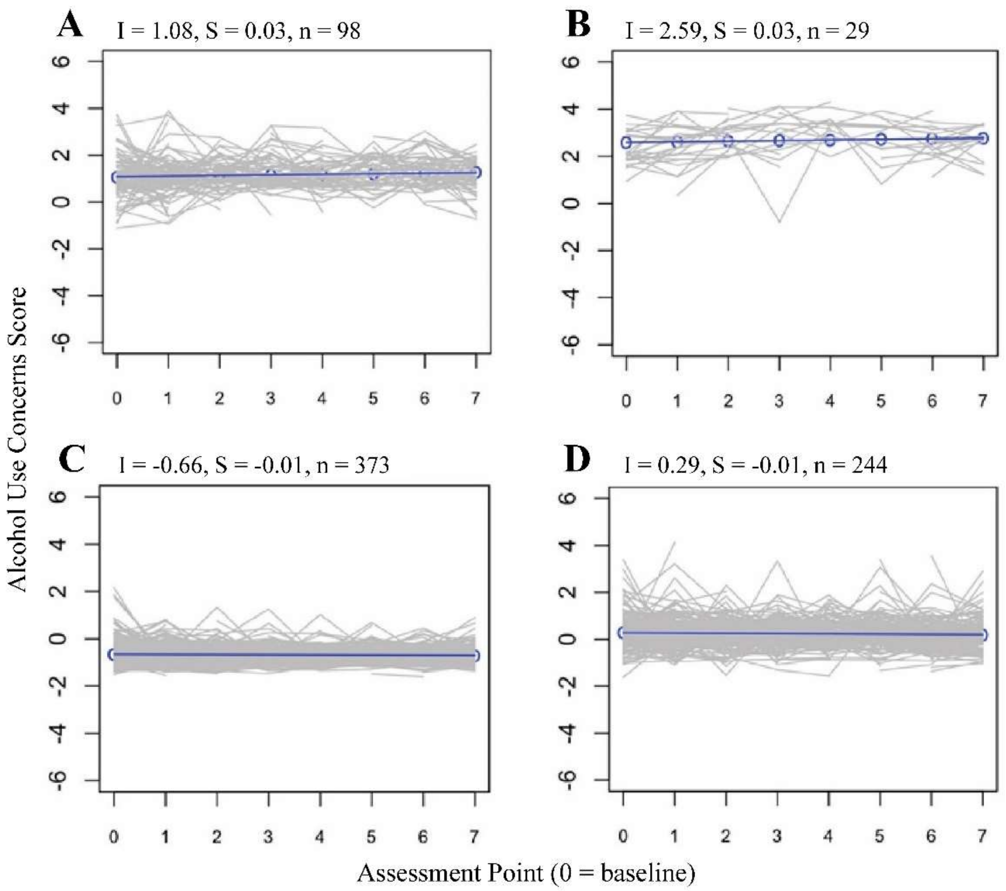

**Figure S3. Growth trajectories on the Alcohol Use Concerns outcome.** Model-estimated growth trajectories in each class are shown by the blue lines, with individual trajectories of participants in the class shaded in grey. Assessment point 0 corresponds to the baseline assessment.

**K-means clustering analysis results**

Several metrics were used to select the optimal number of clusters (i.e., the value of *k*) that reflected participants probabilities of belonging to the worst-faring univariate growth trajectories. Six clusters were identified as optimal across several metrics when the numbers of clusters between $k$ = 2 and $k$ = 10 were evaluated. The ratio of within-cluster sum of squares to the between cluster sum of squares plateaued at $k$ = 6 clusters, such that reductions in this ratio were fairly consistent each additional cluster added to the solution beyond $k$ = 6 clusters and were larger for each increase in clusters prior to $k$ = 6. Internal cluster indices also tended to converge on a 6-cluster solution. The gap statistic compares the observed clustering solution across different values of *k* clusters against solutions obtained with 1000 repeated bootstrap draws of values from simulated data with equivalent variability to one's actual data but no underlying clustering[8]. The resulting gap statistics and associated standard errors are then plotted and the optimal number of clusters is identified based on (a) the extent to which the gap statistic continues to increase after a value of *k* clusters and (b) the overlap in the standard-error of the gap statistic between *k* clusters and $k + 1$ clusters. As displayed in Figure S4, the gap statistic did not show further sharp increases following 6 clusters, and the 95% confidence intervals around the gap statistics associated with 6 and 7 clusters overlapped, suggesting a 6 cluster solution was optimal.

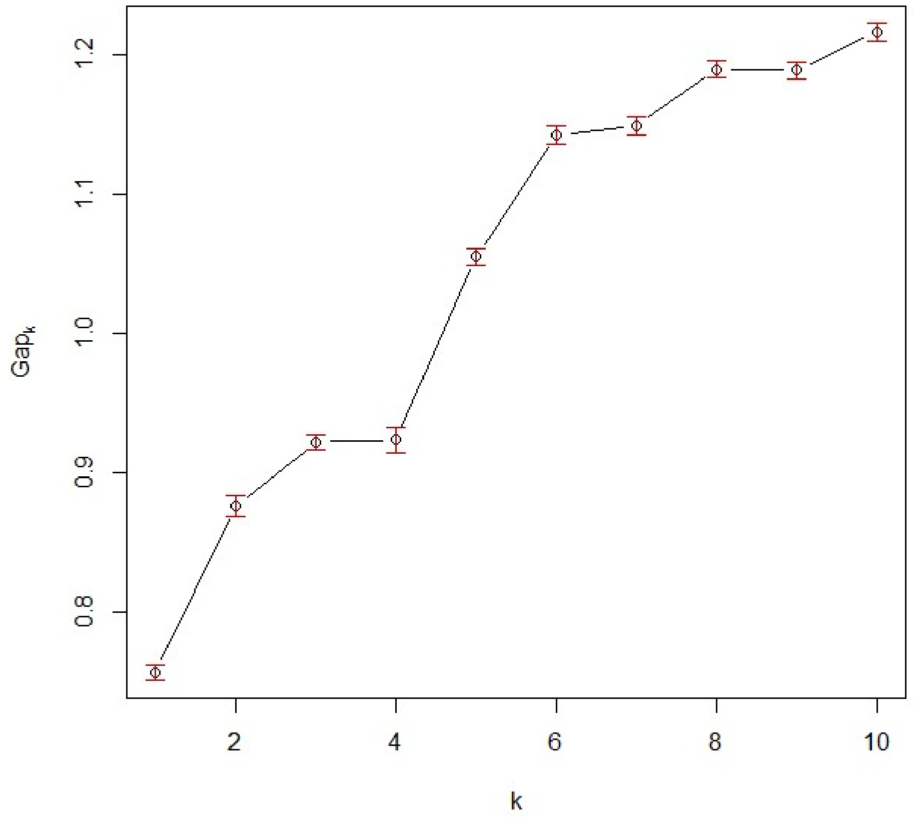

**Figure S4. Plot of gap statistic values across k-means cluster count.** k = number of k-means clusters. $Gap_k$ = value of gap statistic. Points show average gap statistic computed across bootstrap draws and red confidence bands represent the 95% bootstrap confidence intervals around the gap statistic values.

Other internal clustering indices calculated included the average silhouette width, Calinski-Harabasz index, and the Davies-Bouldin index[9–11]. The average silhouette width provides an assessment of the average similarity of participants in each cluster to their assigned cluster as compared with other clusters, with larger values indicating higher similarity. The Calinski-Harabasz index assesses the ratio of between-cluster to within-cluster sums of squares, adjusted by sample size and the number of k-means clusters, with larger values indicating a higher ratio of between-cluster variability relative to within-cluster variability. Finally, the Davies-Bouldin index is computed as a function of the centroid diameters of clusters relative to the distances between clusters, and as such penalizes for the inclusion of very wide clusters spanning a range of possible values. Lower values on the Davies-Bouldin index are indicative of tighter clustering.

The average silhouette width and Davies-Bouldin indexes both agreed with the ratio of within-cluster to between-cluster sums of squares and the gap statistic in identifying 6 clusters as the best solution, whereas the Calinski-Harabasz index indicated two clusters was optimal. In prior experimental comparisons among internal cluster validity indices, the Calinski-Harabasz index has shown a more steeply declining accuracy with larger values of $k$ as compared with certain other indices (i.e., the Silhouette and sums of squares methods)[11], and the average silhouette width and gap statistics have been identified as among the most accurate in comparisons of clustering indices[8,11]. As such, a solution with $k$ = 6 clusters was selected for interpretation based on consensus across clustering indices. The resulting average probabilities of belonging to the worst-faring classes across univariate outcome trajectories are presented by each of the 6 clusters in Table S3.

**Table S3. Table of average probabilities of belonging to the worst-faring latent growth class on each univariate outcome by k-means cluster assignment.**

| Cluster | PCL | PHQ | Soc. | GH | Qual. | PF | Abs. | Rel. | AUD |
|---|---|---|---|---|---|---|---|---|---|
| 1. Worst-faring across domains (n = 52) | .772 | .740 | .815 | .569 | .908 | .459 | .654 | .715 | .113 |
| 2. Problematic alcohol use (n = 44) | .090 | .031 | .051 | .037 | .243 | .007 | .067 | .111 | .952 |
| 3. Physical impairment (n = 107) | .036 | .012 | .093 | .139 | .305 | .156 | .798 | .489 | .053 |
| 4. Trauma & related concerns (n = 46) | .881 | .442 | .468 | .089 | .350 | .156 | .074 | .034 | .046 |
| 5. Low severity across domains (n = 407) | .006 | .008 | .016 | .018 | .028 | .019 | .036 | .030 | .001 |
| 6. Diminished life quality (n = 87) | .025 | .056 | .139 | .171 | .897 | .050 | .076 | .172 | .010 |

*Note.* Values represent average probabilities of belonging to the worst-faring latent growth class on each outcome measure across participants assigned to each cluster. Cluster names are based on the average probabilities of being assigned to the worst-faring growth classes across univariate outcomes in each cluster. PCL = PTSD Checklist. PHQ = Patient Health Questionnaire. Soc. = SF-36 Social Functioning. GH = SF-36 General health. Qual. = Quality of Life. PF = SF-36 Physical Functioning. Abs. = Absolute Activity. Rel. = Relative Activity. AUD = Alcohol Use Disorders Identification Test – Consumption.

Based on the average probabilities of being assigned to the worst-faring latent growth classes across univariate outcomes, Cluster 1 appeared to reflect participants with the poorest faring trajectories across domains. By contrast, the other clusters identified appeared to reflect groups of participants who fared poorly on one or a few outcomes. Cluster 2 reflected participants with a higher probability of being in the latent growth class characterized by elevated alcohol consumption, Cluster 3 reflected participants with a high probability of being in the worst-faring physical activity classes, Cluster 4 reflected participants with a high probability of being in the worst-faring PTSD latent growth class and moderate probabilities of being in the worst-faring depression and social functioning classes, and Cluster 6 was characterized by participants with a high probability of being in the worst-faring life quality latent growth class. Participants in Cluster 5 appeared to have a low probability of being in any of the worst-faring classes across univariate outcome domains.

**Random forest classifier with feature selection results**

Figure S5 displays the areas under the curve (AUCs) of the top one, two, five, 10, 20, 50, and all features identified in the random forest classifier as predictive of membership in the worst-faring distress/impairment class and k-means cluster at each assessment point. Class and cluster membership were predicted as binary outcomes indicating belonging to the class or cluster. The predictive features included at each assessment point represented all measures taken at that year of the study combined with all measures taken at prior years. The top 20 features were selected for interpretation as they showed comparable AUC values relative to larger numbers of features across assessment points. Further, these features tended to represent a similar group of predictive domains with the inclusion of additional measures at each assessment point (see Table 2 of the main manuscript).

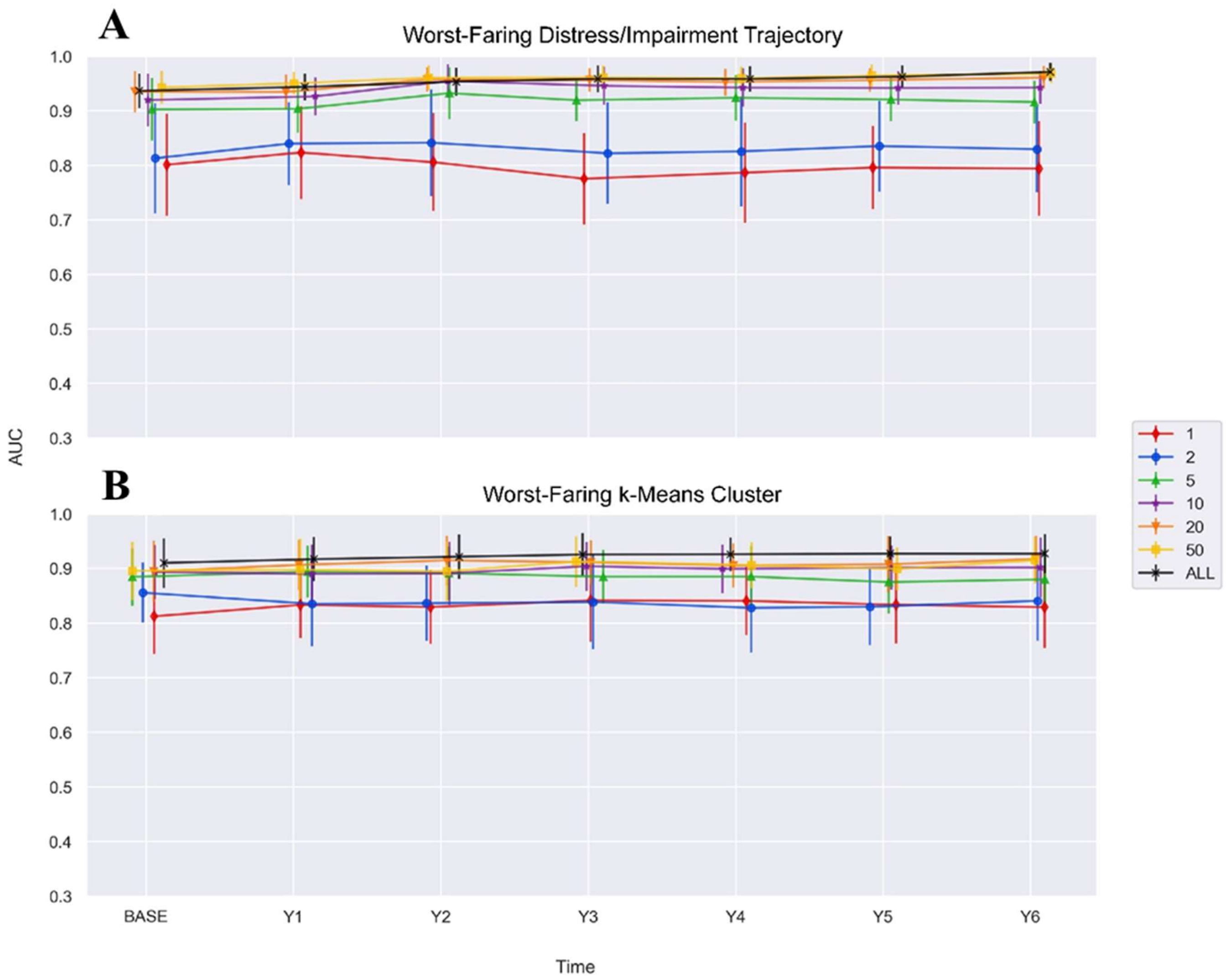

**Figure S5. Plot of areas under the curve for subsets of the top features from the random forest classifier analysis.** AUC = Area under the curve. BASE = Baseline. Y1 - Y7 = Year 1 - Year 7. Lines represent area under the curve computed based on the random forest classifier analysis for the top 1, 2, 5, 10, 20, and 50 features as well as all features. Each time-point includes possible predictors from that time point and all time-points prior. **A**. Areas under the curve predicting membership for the worst-faring distress/impairment class. **B**. Areas under the curve predicting membership for the worst-faring k-means cluster.

#### Supplementary Code

##### Nonlinear Principal Components Analysis SPSS Syntax

```
*Creating split.
USE ALL.
do if $casenum=1.
compute #s_$_1=374.
compute #s_$_2=748.
end if.
do if  #s_$_2 > 0.
compute filter_$=uniform(1)* #s_$_2 < #s_$_1.
compute #s_$_1=#s_$_1 - filter_$.
compute #s_$_2=#s_$_2 - 1.
else.
compute filter_$=0.
end if.
VARIABLE LABELS filter_$ '374 from the first 748 cases (SAMPLE)'.
FORMATS filter_$ (f1.0).
FILTER  BY filter_$.
EXECUTE.
DATASET COPY  Split2.
DATASET ACTIVATE  Split2.
FILTER OFF.
USE ALL.
SELECT IF (filter_$ = 0).
EXECUTE.

*Running NLPCA First Split
DATASET ACTIVATE  DataSet1.
CATPCA VARIABLES=auditcsum  *alcohol use disorders identification test
  mhbhlth  *SF-36 overall health
  mhbpcltotsum *PTSD Checklist
  mhbphys1 *Absolute physical activity rating
  mhbphys5  *Physical activity compared with peers rating
  mhbphysum *SF-36 physical limitations score
  mhbqlty *Quality of life rating
  mhbsocial *SF-36 Social functioning
  phqsum *Patient health questionnaire - depression
  /ANALYSIS=auditcsum(WEIGHT=1,LEVEL=ORDI)
  mhbhlth(WEIGHT=1,LEVEL=ORDI)
  mhbpcltotsum(WEIGHT=1,LEVEL=NUME)
  mhbphys1(WEIGHT=1,LEVEL=ORDI)
  mhbphys5(WEIGHT=1,LEVEL=ORDI)
```

```
  mhbphysum(WEIGHT=1,LEVEL=NUME)
  mhbqlty(WEIGHT=1,LEVEL=ORDI)
  mhbsocial(WEIGHT=1,LEVEL=ORDI)
  phqsum(WEIGHT=1,LEVEL=NUME)
  /DISCRETIZATION=auditcsum(RANKING)
  mhbhlth(RANKING)
  mhbpcltotsum(MULTIPLYING)
  mhbphys1(RANKING)
  mhbphys5(RANKING)
  mhbphysum(MULTIPLYING)
  mhbqlty(RANKING)
  mhbsocial(RANKING)
  phqsum(MULTIPLYING)
  /MISSING=auditcsum(LISTWISE)
  mhbhlth(LISTWISE)
  mhbpcltotsum(LISTWISE)
  mhbphys1(LISTWISE)
  mhbphys5(LISTWISE)
  mhbphysum(LISTWISE)
  mhbqlty(LISTWISE)
  mhbsocial(LISTWISE)
  phqsum(LISTWISE)
  /DIMENSION=3
  /NORMALIZATION=VPRINCIPAL
  /MAXITER=100
  /CRITITER=.00001
  /ROTATION=NOROTATE
  /RESAMPLE=NONE
  /PRINT=LOADING(NOSORT) VAF
  /PLOT=OBJECT(20)
  /SAVE=OBJECT.

*Running NLPCA Second Split
DATASET ACTIVATE  Split2.
CATPCA VARIABLES=auditcsum
  mhbhlth
  mhbpcltotsum
  mhbphys1
  mhbphys5
  mhbphysum
  mhbqlty
  mhbsocial
  phqsum
  /ANALYSIS=auditcsum(WEIGHT=1,LEVEL=ORDI)
```

```
  mhbhlth(WEIGHT=1,LEVEL=ORDI)
  mhbpcltotsum(WEIGHT=1,LEVEL=NUME)
  mhbphys1(WEIGHT=1,LEVEL=ORDI)
  mhbphys5(WEIGHT=1,LEVEL=ORDI)
  mhbphysum(WEIGHT=1,LEVEL=NUME)
  mhbqlty(WEIGHT=1,LEVEL=ORDI)
  mhbsocial(WEIGHT=1,LEVEL=ORDI)
  phqsum(WEIGHT=1,LEVEL=NUME)
  /DISCRETIZATION=auditcsum(RANKING)
  mhbhlth(RANKING)
  mhbpcltotsum(MULTIPLYING)
  mhbphys1(RANKING)
  mhbphys5(RANKING)
  mhbphysum(MULTIPLYING)
  mhbqlty(RANKING)
  mhbsocial(RANKING)
  phqsum(MULTIPLYING)
  /MISSING=auditcsum(LISTWISE)
  mhbhlth(LISTWISE)
  mhbpcltotsum(LISTWISE)
  mhbphys1(LISTWISE)
  mhbphys5(LISTWISE)
  mhbphysum(LISTWISE)
  mhbqlty(LISTWISE)
  mhbsocial(LISTWISE)
  phqsum(LISTWISE)
  /DIMENSION=3
  /NORMALIZATION=VPRINCIPAL
  /MAXITER=100
  /CRITITER=.00001
  /ROTATION=NOROTATE
  /RESAMPLE=NONE
  /PRINT=LOADING(NOSORT) VAF
  /PLOT=OBJECT(20)
  /SAVE=OBJECT.

*Running with full sample
DATASET ACTIVATE  DataSet1.
USE ALL.
CATPCA VARIABLES=auditcsum
  mhbhlth
  mhbpcltotsum
  mhbphys1
  mhbphys5
```

```
mhbphysum
mhbqlty
mhbsocial
phqsum
/ANALYSIS=auditcsum(WEIGHT=1,LEVEL=ORDI)
mhbhlth(WEIGHT=1,LEVEL=ORDI)
mhbpcltotsum(WEIGHT=1,LEVEL=NUME)
mhbphys1(WEIGHT=1,LEVEL=ORDI)
mhbphys5(WEIGHT=1,LEVEL=ORDI)
mhbphysum(WEIGHT=1,LEVEL=NUME)
mhbqlty(WEIGHT=1,LEVEL=ORDI)
mhbsocial(WEIGHT=1,LEVEL=ORDI)
phqsum(WEIGHT=1,LEVEL=NUME)
/DISCRETIZATION=auditcsum(RANKING)
mhbhlth(RANKING)
mhbpcltotsum(MULTIPLYING)
mhbphys1(RANKING)
mhbphys5(RANKING)
mhbphysum(MULTIPLYING)
mhbqlty(RANKING)
mhbsocial(RANKING)
phqsum(MULTIPLYING)
/MISSING=auditcsum(LISTWISE)
mhbhlth(LISTWISE)
mhbpcltotsum(LISTWISE)
mhbphys1(LISTWISE)
mhbphys5(LISTWISE)
mhbphysum(LISTWISE)
mhbqlty(LISTWISE)
mhbsocial(LISTWISE)
phqsum(LISTWISE)
/DIMENSION=3
/NORMALIZATION=VPRINCIPAL
/MAXITER=100
/CRITITER=.00001
/ROTATION=NOROTATE
/RESAMPLE=NONE
/PRINT=LOADING(NOSORT) VAF
/PLOT=OBJECT(20)
/SAVE=OBJECT.
```

**************************************

**Multigroup Confirmatory Factor Analysis R Syntax**

```
#Loading required packages
library(lavaan)
MYH_Data  #Dataset in wide format
#Renaming & scaling measures at each time-point (mhb = baseline, mh1 = 1 year, mh2 = 2-year, (…),
mh7 = 7-year #follow-ups)
MYH_Data$audit_1 <- MYH_Data$auditcsum
MYH_Data$hlth_1 <- MYH_Data$mhbhlth
MYH_Data$physa_1 <- MYH_Data$mhbphys1
MYH_Data$physb_1 <- MYH_Data$mhbphys5
MYH_Data$qlty_1 <- MYH_Data$mhbqlty
MYH_Data$soc_1 <- MYH_Data$mhbsocial
MYH_Data$phq_1 <- MYH_Data$phqsum
MYH_Data$pcltot_1 <- MYH_Data$mhbpcltotsum/10
MYH_Data$physum_1 <- MYH_Data$mhbphysum/10

MYH_Data$audit_2 <- MYH_Data$mh1auditsum
MYH_Data$hlth_2 <- MYH_Data$mh1hlth
MYH_Data$physa_2 <- MYH_Data$mh1phys1
MYH_Data$physb_2 <- MYH_Data$mh1phys5
MYH_Data$qlty_2 <- MYH_Data$mh1qlty
MYH_Data$soc_2 <- MYH_Data$mh1social
MYH_Data$phq_2 <- MYH_Data$mh1ppqsum
MYH_Data$pcltot_2 <- MYH_Data$mh1pclsum/10
MYH_Data$physum_2 <- MYH_Data$mh1physum/10

MYH_Data$audit_3 <- MYH_Data$mh2auditsum
MYH_Data$hlth_3 <- MYH_Data$mh2hlth
MYH_Data$physa_3 <- MYH_Data$mh2phys1
MYH_Data$physb_3 <- MYH_Data$mh2phys5
MYH_Data$qlty_3 <- MYH_Data$mh2qlty
MYH_Data$soc_3 <- MYH_Data$mh2social
MYH_Data$phq_3 <- MYH_Data$mh2ppqsum
MYH_Data$pcltot_3 <- MYH_Data$mh2pclsum/10
MYH_Data$physum_3 <- MYH_Data$mh2physum/10

MYH_Data$audit_4 <- MYH_Data$mh3auditsum
MYH_Data$hlth_4 <- MYH_Data$mh3hlth
MYH_Data$physa_4 <- MYH_Data$mh3phys1
MYH_Data$physb_4 <- MYH_Data$mh3phys5
MYH_Data$qlty_4 <- MYH_Data$mh3qlty
MYH_Data$soc_4 <- MYH_Data$mh3social
MYH_Data$phq_4 <- MYH_Data$mh3ppqsum
```

```
MYH_Data$pcltot_4 <- MYH_Data$mh3pclsum/10
MYH_Data$physum_4 <- MYH_Data$mh3physum/10

MYH_Data$audit_5 <- MYH_Data$auditsum
MYH_Data$hlth_5 <- MYH_Data$hlth
MYH_Data$physa_5 <- MYH_Data$phys1
MYH_Data$physb_5 <- MYH_Data$phys5
MYH_Data$qlty_5 <- MYH_Data$qlty
MYH_Data$soc_5 <- MYH_Data$social
MYH_Data$phq_5 <- MYH_Data$mh4phqsum
MYH_Data$pcltot_5 <- MYH_Data$mh4pclsum/10
MYH_Data$physum_5 <- MYH_Data$physum/10

MYH_Data$audit_6 <- MYH_Data$mh5auditsum
MYH_Data$hlth_6 <- MYH_Data$mh5hlth
MYH_Data$physa_6 <- MYH_Data$mh5phys1
MYH_Data$physb_6 <- MYH_Data$mh5phys5
MYH_Data$qlty_6 <- MYH_Data$mh5qlty
MYH_Data$soc_6 <- MYH_Data$mh5social
MYH_Data$phq_6 <- MYH_Data$mh5ppqsum
MYH_Data$pcltot_6 <- MYH_Data$mh5pclsum/10
MYH_Data$physum_6 <- MYH_Data$mh5physum/10

MYH_Data$audit_7 <- MYH_Data$mh6auditsum
MYH_Data$hlth_7 <- MYH_Data$mh6hlth
MYH_Data$physa_7 <- MYH_Data$mh6phys1
MYH_Data$physb_7 <- MYH_Data$mh6phys5
MYH_Data$qlty_7 <- MYH_Data$mh6qlty
MYH_Data$soc_7 <- MYH_Data$mh6social
MYH_Data$phq_7 <- MYH_Data$mh6ppqsum
MYH_Data$pcltot_7 <- MYH_Data$mh6pclsum/10
MYH_Data$physum_7 <- MYH_Data$mh6physum/10

MYH_Data$audit_8 <- MYH_Data$mh7auditsum
MYH_Data$hlth_8 <- MYH_Data$mh7hlth
MYH_Data$physa_8 <- MYH_Data$mh7phys1
MYH_Data$physb_8 <- MYH_Data$mh7phys5
MYH_Data$qlty_8 <- MYH_Data$mh7qlty
MYH_Data$soc_8 <- MYH_Data$mh7social
MYH_Data$phq_8 <- MYH_Data$mh7ppqsum
MYH_Data$pcltot_8 <- MYH_Data$mh7pclsum/10
MYH_Data$physum_8 <- MYH_Data$mh7physum/10

##Converting Wide data to Long
```

```
A <- match("audit_1", names(MYH_Data))
B <- match("physum_8", names(MYH_Data))
C <- match("physum_8",names(MYH_Data[,c(1,A:B)]))
SubData <- MYH_Data[,c(1,A:B)]
MYH_Long <- reshape(SubData,varying = c(2:C),
timevar = "time", idvar = "ParticipantID",
direction = "long", sep = "_")
Data <- as.data.frame(MYH_Long, head=T)

#Identifying variables to be estimated with ordinal scaling
Ordvars <- c(“audit”,"physa","physb","hlth","qlty","soc")

####Bifactor Model, one group
CF_1g_2f <- "
#measurement model
ComFact_B =~ hlth + pcltot + physa + physb + physum +
qlty + soc + phq
+ 0*audit
PhysFact_B =~ pcltot + physa + physb + phq
+ 0*audit
#Factor variance/covariance
ComFact_B~~1*ComFact_B
PhysFact_B~~1*PhysFact_B
ComFact_B ~~ 0*PhysFact_B"

CF_1g_2f_fit <- cfa(CF_1g_2f, data=MYH_Long,
ordered = Ordvars, std.lv = TRUE)
#summary(CF_1g_2f_fit, standardized = T)
fitMeasures(CF_1g_2f_fit, c("cfi","rmsea","srmr"))

####Bifactor Model, multi-group, free loadings & int's
CF_Mg_2f <- "
#measurement model
ComFact_B =~ hlth + pcltot + physa + physb + physum +
qlty + soc + phq
+ 0*audit
PhysFact_B =~ pcltot + physa + physb + phq
+ 0*audit
#Factor variance/covariance
ComFact_B~~1*ComFact_B
PhysFact_B~~1*PhysFact_B
ComFact_B ~~ 0*PhysFact_B"

CF_Mg_2f_fit <- cfa(CF_Mg_2f, data=MYH_Long,
```

```
ordered = Ordvars, std.lv = TRUE, group = "time")
#summary(CF_Mg_2f_fit, standardized = T)
fitMeasures(CF_Mg_2f_fit, c("cfi","rmsea","srmr"))

####Bifactor Model, multi-group, fixed loadings & free int's
CF_Fl_2f <- "
#measurement model
ComFact_B =~ hlth + pcltot + physa + physb + physum +
qlty + soc + phq
+ 0*audit
PhysFact_B =~ pcltot + physa + physb + phq
+ 0*audit
#Factor variance/covariance
ComFact_B~~1*ComFact_B
PhysFact_B~~1*PhysFact_B
ComFact_B ~~ 0*PhysFact_B"

CF_Fl_2f_fit <- cfa(CF_Fl_2f, data=MYH_Long,
ordered = Ordvars, std.lv = TRUE, group = "time",
group.equal = c("loadings"))
#summary(CF_Fl_2f_fit, standardized = T)
fitMeasures(CF_Fl_2f_fit, c("cfi","rmsea","srmr"))

####Bifactor Model, multi-group, fixed loadings & int's
CF_Fi_2f <- "
#measurement model
ComFact_B =~ hlth + pcltot + physa + physb + physum +
qlty + soc + phq
+ 0*audit
PhysFact_B =~ pcltot + physa + physb + phq
+ 0*audit
#Factor variance/covariance
ComFact_B~~1*ComFact_B
PhysFact_B~~1*PhysFact_B
ComFact_B ~~ 0*PhysFact_B"

CF_Fi_2f_fit <- cfa(CF_Fi_2f, data=MYH_Long,
ordered = Ordvars, std.lv = TRUE, group = "time",
group.equal = c("loadings","intercepts"))
#summary(CF_Fi_2f_fit, standardized = T)
fitMeasures(CF_Fi_2f_fit, c("cfi","rmsea","srmr"))

####Bifactor Model, multi-group, fixed loadings & res's
CF_Fr_2f <- "
```

```
#measurement model
ComFact_B =~ hlth + pcltot + physa + physb + physum +
qlty + soc + phq
+ 0*audit
PhysFact_B =~ pcltot + physa + physb + phq
+ 0*audit
#Factor variance/covariance
ComFact_B~~1*ComFact_B
PhysFact_B~~1*PhysFact_B
ComFact_B ~~ 0*PhysFact_B"

CF_Fr_2f_fit <- cfa(CF_Fr_2f, data=MYH_Long,
ordered = Ordvars, std.lv = TRUE, group = "time",
group.equal = c("loadings","residuals"))
#summary(CF_Fr_2f_fit, standardized = T)
fitMeasures(CF_Fr_2f_fit, c("cfi","rmsea","srmr"))

####Bifactor Model, multi-group, fixed loadings, int's, resids
CF_Fall_2f <- "
#measurement model
ComFact_B =~ hlth + pcltot + physa + physb + physum +
qlty + soc + phq
+ 0*audit
PhysFact_B =~ pcltot + physa + physb + phq
+ 0*audit
#Factor variance/covariance
ComFact_B~~1*ComFact_B
PhysFact_B~~1*PhysFact_B
ComFact_B ~~ 0*PhysFact_B"

CF_Fall_2f_fit <- cfa(CF_Fall_2f, data=MYH_Long,
ordered = Ordvars, std.lv = TRUE, group = "time",
group.equal = c("loadings","intercepts","residuals"))
#summary(CF_Fall_2f_fit, standardized = T)
fitMeasures(CF_Fall_2f_fit, c("cfi","rmsea","srmr"))

##Comparing models at each level of constraint##
anova(CF_Mg_2f_fit,CF_Fl_2f_fit) #Comparing models with free parameters across groups and fixed
loadings
anova(CF_Mg_2f_fit,CF_Fi_2f_fit) #Comparing models with free parameters across groups and fixed
loadings/intercepts
anova(CF_Mg_2f_fit,CF_Fr_2f_fit) #Comparing models with free parameters across groups and fixed
loadings/residuals
```

```
library(semPlot)
Bifactor_Bestfit <- semPaths(CF_Fr_2f_fit, what = "stand",
layout = "tree3", residuals = T,intercepts = F,subRes = 360,
panelGroups = F, asize = 2, edge.color = "blue",
fade = T, esize = 2, asize = 2)
```

**Python Code for Latent Class Growth Analysis (LCGA) Mplus Automation**

```
@author: kirsh012

Description:
    This file will automatically generate MPlus input files based on the
    parameters you choose.
"""

import numpy as np
import csv
import string
#np.set_printoptions(precision=2, suppress = True)

# Save to path
savepath = 'MYH'

# Define the name of the file
scriptname = "MYH_c5_ppq_tom"

# The title of your script
title = "MYH_c5_ppq_tom" # "LCGA_WS_c1_TEST"

# Specify the path to the data file to run your model on
filepath =  "\"/Users/kirsh012/Box/MYH Study/MYH_ppq_data.dat\""
# The names of the columns in your dataset
cols = []
with open("/Users/kirsh012/Box/MYH Study/MYH_ppq_columns.txt", 'r') as f:
    reader = csv.reader(f, delimiter = '\t')
    for row in reader:
        cols.append(row)
columns = cols[0]

# The outcome variables in your dataset
out_cols = []
with open("/Users/kirsh012/Box/MYH Study/MYH_ppq_outcome_vars.txt", 'r') as fo:
    reader = csv.reader(fo, delimiter = '\t')
    for row in reader:
        out_cols.append(row)
outcome_cols = out_cols[0]

# Specify what value your missing values are represented by
missing_vals = "*"
```

```
# The number of latent classes
num_classes = "5"

# Which algorithm Mplus should use
model_type = "MIXTURE" # Add COMPLEX for RAISE data

# Number of random sets of starting values (default = 10)
num_rand_start = "200"
# Number of final optimizations (default = 2)
num_opt = "20"
# Number of highset loglikelihood estimations after optimization
iterations = "10"

# Create array of time intervals for your outcome variables
time = np.array([0, 1, 2, 3, 4, 5, 6, 7])#np.sqrt(np.linspace(0, 24, 5))

# Specify model and free estimates across all sets
model = "OVERALL"

# Tells the script to set the intercept and slope variance to 0 if True
latent = True

# Tells the script to include the Residuals output
residuals = True

# The different tech variables to include
tech_num = [1, 8, 11, 14, 4]

# Tells script to output plots if plot is True
plot = True
plot_type = "PLOT3"

# Formats times to strings and concatenates into one string
time = [str(x)[:4] for x in time]
time_intervals = " ".join(time)

# Splits strings on white spaces and converts to list
#columns = columns.split() # Comment out when uploading column variables
oc = outcome_cols#.split() # Comment out split when uploading outcome variables
ti = time_intervals.split()

# Join the outcome cols and time intervals to match
factors = " ".join([f"{var}@{intr}" for var, intr in zip(oc, ti)])
tech = " ".join([f"tech{i}" for i in tech_num])
```

```
final_columns = [columns[x:x+4] for x in range(0, len(columns), 4)] #4
final_outcome_cols = [outcome_cols[x:x+4] for x in range(0, len(outcome_cols), 4)] #4
final_factors = [factors.split()[x:x+3] for x in range(0, len(factors.split()), 3)] #3

with open(f'trajectory_{savepath}/{scriptname}.inp', 'w') as script:

    script.write(f"Title: {title}\n")

    script.write(f"Data: file is {filepath};\n")
    numcol = 0
    for group in final_columns:
        group = str(group)
        group = group.translate(str.maketrans('', '', string.punctuation))
        #group = re.sub(r'\s', r'\t', group)
        if numcol == 0:
            if len(final_columns) != 1:
                script.write(f"Variable: names are {group}\n")
            else:
                script.write(f"Variable: names are {group};\n")

        elif numcol != (len(final_columns) - 1):
            script.write(f"\t\t{group}\n")
        else:
            script.write(f"\t\t{group};\n")
        numcol += 1

    outnum = 0
    for groupset in final_outcome_cols:
        groupset = str(groupset)
        groupset = groupset.translate(str.maketrans('', '', string.punctuation))
        #groupset = re.sub(r'\s', r'\t', groupset)
        if outnum == 0:
            if len(final_outcome_cols) != 1:
                script.write(f"\tusevar= {groupset}\n")
            else:
                script.write(f"\tusevar= {groupset};\n")

        elif outnum != (len(final_outcome_cols) - 1):
            script.write(f"\t\t{groupset}\n")
        else:
            script.write(f"\t\t{groupset};\n")
        outnum += 1
    if effect:
```

```
    script.write(f"\tcluster={hospital_effect};\n")

script.write(f"\tmissing={missing_vals};\n")
script.write(f"\tCLASSES=c({num_classes});\n")

script.write(f"Analysis: type={model_type};\n")
script.write(f"\tSTARTS={num_rand_start} {num_opt};\n")
script.write(f"\tSTITERATIONS={iterations};\n\n")

script.write(f"Model: %{model}%\n")
facnum = 0
for factor in final_factors:
    factor = str(factor)
    factor = factor.translate(str.maketrans('', '', r"\[\]\',"))
    if facnum == 0:
        if len(final_factors) != 1:
            script.write(f"\ti s|{factor}\n")
        else:
            script.write(f"\ti s|{factor};\n")

    elif facnum != (len(final_factors) - 1):
        script.write(f"\t    {factor}\n")
    else:
        script.write(f"\t    {factor};\n")
    facnum += 1

if latent:
    script.write(f"\ti-s@0;\n\n")

if residuals:
    output = f"Output: residual sampstat standardized {tech};\n"
    script.write(output)
else:
    script.write(f"Output: sampstat standardized {tech};\n")
if plot:
    snum = 0
    for serie in final_outcome_cols:
        serie = str(serie)
        serie = serie.translate(str.maketrans('', '', string.punctuation))
        if snum == 0:
            if len(final_outcome_cols) != 1:
                script.write(f"PLOT: SERIES={serie}\n")
            else:
                script.write(f"PLOT: SERIES={serie} (s);\n")
```

```
    elif snum != (len(final_outcome_cols) - 1):
        script.write(f"\t{serie}\n")
    else:
        script.write(f"\t{serie} (s);\n")

    snum += 1
script.write(f"\tTYPE={plot_type};\n")
```

**R Syntax for K-Means Clustering of LCGA Class Probabilities**

```
#Loading required packages and data
library(cluster)
library(clusterCrit)
library(factoextra)
Probs <- as.data.frame(read.csv("bad_class_probabilities.csv", head=T)) #dataframe with worst-faring
class probabilities
Clean_Probs <- as.data.frame(na.omit(Probs[,2:10])) #selecting cases with class probabilities for the
worst-faring classes
set.seed(2847)

####Selecting best-fitting model across 25 possible models for each value of k from 2-10####
#Running k-means multiple times for each cluster number from 2-10 clusters (stored in Modlist)
Outs1 <- as.data.frame(matrix(nrow = 25, ncol = 10))
Outs2 <- as.data.frame(matrix(nrow = 25, ncol = 10))
Modlist <- vector(mode = "list", length = 250)
for (i in 2:10){
 for (j in 1:25){
 d <- kmeans(Clean_Probs,i, iter.max = 100,
 nstart = 5)
 Outs[j,(i-1)] <- d$betweenss/(d$betweenss+d$tot.withinss)
 Modlist[[((i-2)*25)+j]] <- d
 }}

#Finding optimal model for each number of clusters based on the ratio
#of between sum of squares to total sum of squares for each number of clusters (stored in Optim)
Optim <-c(rep(NA,9))
for (i in 1:9){
 num <- which.max(Outs[,i])
 Optim[i] <- num + (25*(i-1))
}

####Finding best number of clusters, based on optimal models selected in previous step####
#Between vs Total Ratio (decrements in slope at 3, 4, and 6 clusters)
OptFinal <- c(rep(NA,9))
for (i in 1:9){
 Mnum <- Optim[i]
 b <- Modlist[[Mnum]]
 v <- b$betweenss/(b$betweenss+b$tot.withinss)
 OptFinal[i] <- v
 }
plot(x = c(2:10), y = OptFinal, type = "o",
 xlab = "K", ylab = "BetweenSS/TotalSS")
```

```
#Silhouette Widths (suggests 2 or 6 clusters)
library(cluster)
SilWidths <- c(rep(NA,9))
for (i in 1:9){
 Mnum <- Optim[i]
 b <- Modlist[[Mnum]]
 s <- silhouette(b$cluster,dist(Clean_Probs))
 SilWidths[i] <- summary(s)$avg.width
 }
plot(x = c(2:10), y = SilWidths, type = "o",
 xlab = "K", ylab = "Avg. Silhouette Width")

#Gap Statistics (suggests 6 clusters)
Gaps <- clusGap(Clean_Probs, FUN = kmeans, K.max = 10, B = 1000)
plot(Gaps)

#Calinski Harabasz Index (suggests 2 clusters)
C_H <- c(rep(NA, 9))
for (i in 1:9){
 Mnum <- Optim1[i]
 b <- Modlist[[Mnum]]
 IntIdx <- intCriteria(as.matrix(Clean_Probs),b$cluster,"all")
 C_H[i] <- IntIdx[["calinski_harabasz"]]
 }
plot(x = c(2:10), y = C_H,type = "o",
    xlab = "K", ylab = "Calinski_Harabasz Index")

#Davies-Bouldin Index (suggests 6 clusters)
D_B <- c(rep(NA, 9))
for (i in 1:9){
 Mnum <- Optim1[i]
 b <- Modlist[[Mnum]]
 IntIdx <- intCriteria(as.matrix(Clean_Probs),b$cluster,"all")
 D_B[i] <- IntIdx[["davies_bouldin"]]
 }
plot(x = c(2:10), y = D_B,type = "o",
    xlab = "K", ylab = "Davies-Bouldin Index")
```
